# Women’s experiences in the food environment and their association with fruit and vegetables intake: Insights from Northern Tanzania

**DOI:** 10.1101/2025.07.01.25330643

**Authors:** Nishmeet Singh, Lilia Bliznashka, Fusta Azupogo, Quinn Marshall, Neha Kumar, Evangelista Malindisa, Kidola Jeremiah, Sonja Y Hess, Joyce L Kinabo, Deanna K. Olney, Lindsay M. Jaacks, Alexandra L. Bellows

## Abstract

**Background:** There is limited information from rural low-income contexts about consumers’ buying behaviours of fruit and vegetables (F&V) and accessibility to F&V in the food environments, which may inform interventions to increase F&V intake.

**Objectives:** We examined how women living in rural northern Tanzania experience the food environments by exploring buying patterns, perceptions, and accessibility of F&V, and their association with women’s F&V intake.

**Methods:** We used cross-sectional data from 2,597 women living in the Arusha and Kilimanjaro regions. Self-reported experiences of the food environments for F&V included buying frequency, sources, availability, and convenience. Accessibility was measured using geospatial measures of distance and time between home and reported typical buying sources. Data from a 30-day F&V food frequency questionnaire were used to calculate two scores that captured intake frequency and variety. We conducted a descriptive analysis of women’s experience indicators and tested their associations with the scores using multivariable and Poisson regression models, controlling for covariates.

**Results:** On average, 5% and 35% of women reported daily buying of F&V, respectively. Fruit was mostly bought from markets: F: 80%, V: 40%. Two-thirds of respondents perceived F&V as available (F: 65%, V: 60%). Median (IQR) distance and time to fruit sources was 9 km (2,19), 39 min (19,78), and to vegetable sources was 3 km (1,10), 32 min (8-69). Compared to women who reported making daily purchases of F&V, those who purchased F&V weekly or monthly reported lower frequency and diversity of F&V intake. Perceptions that F&V were less available and at a longer distance, but not time, were associated with lower frequency and variety of vegetable intake.

**Conclusion:** Buying frequency, perceived availability, and distance to markets were associated with women’s F&V intake frequency and variety, underscoring the need to consider these and other factors in food environments to increase F&V intake.

## Introduction

Fruit and vegetables are vital components of a healthy diet, as they are rich in essential micronutrients and fibre (Asaduzzaman et al., 2018). People who consume fruit and vegetables have lower risks of cardiovascular diseases, cancer, and all-cause mortality (Aune et al., 2017), potentially due to their high fibre content and antioxidant properties (Gill et al., 2021). Locally and seasonally sourced plant-based foods also have some of the lowest environmental footprints compared to other foods (Clark et al., 2019). The World Health Organisation (WHO) recommends daily consumption of at least 400 grams or five servings of combined fruit and vegetables (WHO, 2003). There is limited evidence on the intake of fruit and vegetables in Africa. Available studies suggest that intake has increased over the last decade; however, it still falls short of the WHO recommendation (Mensah et al., 2021). In Africa, the mean per capita daily intake among adults and children is 268 grams, with higher levels observed among individuals living in rural areas (Mensah et al., 2021). In East Africa, low intake of fruit and vegetables is one of the leading dietary risk factors for deaths and disability-adjusted life-years (Afshin et al., 2019).

Food environment and consumer behaviours are considered immediate drivers of food choices and diets that interact dynamically as part of agri-food systems (HLPE, 2020). While there are several definitions of food environment in the literature, we defined it as the interface where people interact with the broader food system to acquire and consume food (Turner et al., 2018). The term ‘interface’ describes the reciprocal relationship between food environments, which are physical or external spaces, and consumer behaviours as shaped by individual, family and sociocultural factors (Ambikapathi et al., 2024; Turner et al., 2018). External food environments are categorised as built (formal and informal sources), natural (wild or cultivated), and kinship and community sources (Bogard et al., 2021; Downs et al., 2020). Over the last decade, several food environment studies, particularly in low-income and transitioning agri-food systems contexts, have primarily focused on the external domain in urban or peri-urban, and/or formal retail settings (Karanja et al., 2022; Turner et al., 2020). In light of this trend, recent calls have been made for the evidence-based measurement of personal experiences or ‘people-centred’ food environment in rural contexts (Downs et al., 2020; Dzanku et al., 2024; Fanzo et al., 2021; Spires et al., 2023).

Reviews from the last five years have identified a dearth of research on drivers of fruit and vegetables choices and their relationship to diets in rural contexts, including Africa (Choudhury et al., 2025; Harris et al., 2022; Karanja et al., 2022). This research gap is pertinent to both the low supply and intake levels of fruits and vegetables, and the rise in the availability and purchasing of highly- and ultra-processed foods in rural areas (Kalmpourtzidou et al., 2020; Reardon et al., 2024; Sauer et al., 2021). These trends can sway food choices towards unhealthy diets. There is a need to better understand consumer experiences in the food environment among rural populations to improve the consumption of fruit and vegetables (Choudhury et al., 2025; Stadlmayr et al., 2023).

In rural Tanzania, dietary intake studies reveal low intakes of fruit and vegetables; however, information is limited on the drivers of food choices, including consumers’ experiences with purchasing fruit and vegetables within their food environment (Amunga et al., 2024). The primary objectives of this paper were: 1) to describe how women living in rural Tanzania experienced the food environment while buying fruit and vegetables, and 2) to examine the association of food environment experiences with women’s reported frequency and variety of fruit and vegetables intake.

## Methods

We analysed cross-sectional data on 2,597 women aged 15 to 49 years living in 33 villages across five districts in the Arusha and Kilimanjaro regions of northern Tanzania **(supplement figure 1).** We used food environment data from 1,260 food vendors and 15 markets located in and around the selected villages to assess spatial accessibility. The women and food environment data were collected as part of the baseline survey for the ‘Fruit and Vegetables for Sustainable Healthy Diets’ (FRESH) end-to-end evaluation conducted between October 2023 and January 2024. The evaluation aims to evaluate the impact of different combinations of supply, demand and food environment interventions on women’s fruit and vegetables intake and vegetable production (Bliznashka et al., 2023).

Adult women were enrolled in the baseline survey if they were living in the study area, were aged 15 to 49 years, had a biological child (adolescent aged 10 to 14 years), and planned to continue residing in the same area for the next 12 months. An initial list of eligible households was obtained from village leaders, and eligible women were randomly invited to participate in the study until the field team reached the necessary sample size.

### Data Collection

To understand women’s experiences in the food environment, we designed a module on drivers of food choices that encompassed women’s self-reported buying patterns and personal perceptions separately for fruit and vegetables for the previous 30 days, based on exiting literature (Gupta et al., 2023; Green and Glanz 2015, Yamaguchi 2022, Lo 2019, Gao 2022). The buying patterns and perceptions questions were administered to a subsample of women who reported buying any fruit or vegetables in the previous 30 days, and only for the primary source of fruit and the primary source of vegetables **(supplement table 1).** To assess buying patterns, women were asked about the typical sources from which they purchased (ranking the top three), the frequency of buying, the origin of their trips (home, work, or farm), and their primary mode of transport (car, public transport, walk/cycle, etc.). The primary source was identified as the source women ranked highest among those from which they typically bought most of their fruit or vegetables. The questions related to perceptions covered availability (year-round variety at the desired level at the primary source and the presence of a market in the village for fruit or vegetables) and the convenience of travelling to the primary source, considering travel time, childcare responsibilities, and household chores. To further understand some of the reported experiences, we asked women to describe their reasons for selecting the primary source and the inconvenience of accessibility using open-ended questions.

Intake data were collected using a 30-day non-quantitative food frequency questionnaire (FFQ) for fruits and vegetables. The FFQ included a list of 35 fruit and 31 vegetables, with the option to report additional items. For each fruit and vegetables question, the response options ranged from “2-3 times per day” to “never”. The fruit and vegetables lists were adapted from existing literature (Lukmanji et al., 2013; Zack et al., 2018) and further revised during the enumerators’ training and after the pilot surveys.

Using an enumerator-based food environment census questionnaire, we collected data on the geolocation of all food outlets and markets in and around the study villages, and whether outlets sold fruit, vegetables, and other food groups. This assessment covered 1,260 fruit and vegetables outlets in the villages, including kiosks (street stalls) and retail shops, and 15 main fruit and vegetables markets outside the village boundaries identified by community leaders as accessed by community members.

Covariate data **(supplement table 2)** were collected using a structured household questionnaire and included demographic data on key household characteristics (wealth, food insecurity, size, dependency ratio, consumption from own production, and geo-coordinates) as well as for the targeted women (age, education, marital status, and primary occupation).

### Exposure variables

Exposure variables were created separately for fruit and vegetables and classified as perceived or objective measures (Toure et al., 2021).

Perceived measures of buying fruit and vegetables from the primary source included frequency of buying (daily/weekly/monthly), year-round availability (mostly/sometimes/rarely), presence of a market in the village (yes/no), and convenience (easy/neutral/difficult).

Objective measures included spatial accessibility, which was estimated using two Geographic Information System (GIS)-based proximity indicators between the household location and the reported typical sources for fruit and vegetables: distance and time (Charreire et al., 2010). Calculating accessibility from household to sources is a commonly used method in the food environment literature (Choudhury et al., 2025). These indicators were calculated using Google Maps Direction API in R software (Dorman et al., 2023; Posit Team, 2025). Both indicators were calculated based on the shortest network path between home and the sources, conditional on the mode of transport reported, such as driving, walking, or public transport (Thornton et al., 2017). For each household, the typical source was identified as the primary source of fruit and vegetables reported by the women in the household survey. However, when a non-market source was reported as the primary source (e.g., shop or kiosk), the geographically nearest source to the household was used, due to our inability to match the non-standardised names of non-market outlets reported by the women. More information about estimating accessibility using geospatial data, including information on data cleaning and adjustments, is available in Supplementary Text 1. We reported all estimates for distance and time as round-trip journeys. For example, a one-sided estimate of 1 km and 30 mins from the Google Maps API was doubled to 2 km and 60 mins to arrive at the round-trip journey. **Figure 1** depicts our method for a sample context.

**Figure 1.**
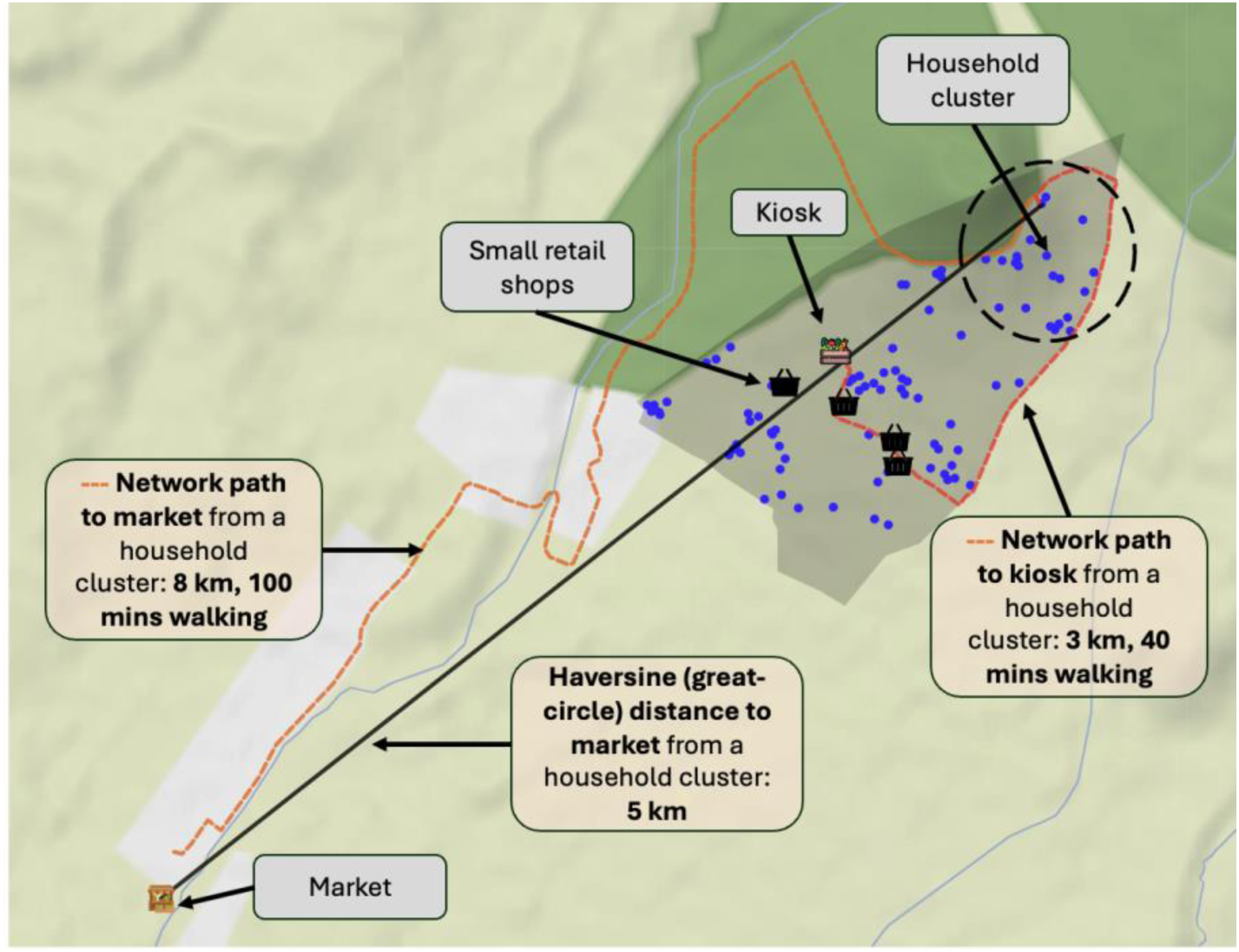
Map of a sample context describing the accessibility method used in the study. Notes: The map shows household clusters (in blue), fruit and vegetable sources (market, kiosks and retail shops) and travel routes from a sample context. It depicts the distance, and time estimates from the ‘network path’ method and the ‘Haversine (great-circle)’ method between a household cluster and sources. The path methods represent the shortest travel distance and time, taking into account the roads and available walkways. The Haversine distance method calculates distance between two points over the Earth’s surface (see (Gamba et al., 2015) for a systematic review of geospatial methods in the food environment).

## Outcome variables

Fruit and vegetables intake was estimated based on the frequency and variety of consumption. We calculated an intake frequency score separately for fruit and vegetables by assigning daily weights to each choice option in the FFQ and then summing the reported consumption values (CDC: Centers for Disease Control and Prevention, 2005; Siebelink et al., 2011) (**supplement text 2**). The maximum possible score was 90 for fruit and 80 for vegetables. A higher score indicated a greater frequency of fruit and vegetables intake. In the descriptive analysis, we reported the proportion of women consuming any fruit or vegetables daily by combining the three choice options in the FFQ: “2-3 times per day”, “once per day,” and “5-6 times a week”.

To estimate the variety of fruit and vegetables intake (number of different fruit and vegetable items within a food group), we used the Global Dietary Recommendation (GDR) score. We calculated a separate daily fruit-GDR score and a daily vegetable GDR score as a sum of three fruit and vegetables food groups consumed (range 0-3) (Pastori et al., 2023). The included food groups were vitamin A-rich fruit, citrus fruit, and other fruit; and vitamin A-rich vegetables, dark green leafy vegetables, and other vegetables. Each fruit and vegetables item was categorised either based on the Codex Alimentarius definition or, otherwise, the FAO’s Minimum Diet Diversity (MDD) guidelines for women (Codex Alimentarius definition used in Arimond et al., 2010; FAO, 2021) **(supplement text 3).** A higher fruit or vegetable GDR score indicated a greater variety of intake.

### Statistical analysis

We calculated means, standard deviations, and frequencies for household and women’s demographic characteristics, and for indicators of women’s experiences in the food environment. We also presented the proportion of women consuming any fruit and vegetables daily and the proportion of fruit and vegetables items consumed in the previous 30 days. A food insecurity score was calculated and reported as the proportion of households with any food insecurity (scores of 1-8), mild food insecurity (scores of 1-3), and moderate or severe food insecurity (scores of 4-8) (Singh et al., 2023).

We used regression models, controlling for covariates and adjusting for clustering by village using robust standard errors, to estimate the associations between outcome and exposure variables, separately for fruit and vegetables. We ran separate regressions for four sets of exposure variables: 1) buying frequency; 2) buying perceptions regarding availability throughout the year, market in the village, and convenience; 3) round trip journey distance between home and source, quintiles; and 4) round trip journey time between home and source, quintiles. Covariates varied for each model based on associations with the outcome and individual exposure variables in univariate models at P<0.20. The initial set of covariates comprised of respondent age, education, occupation, marital status, primary occupation, household size, household dependency ratio, household wealth (calculated using a principal component analysis from 12 assets and seven housing characteristics), any household food insecurity, and a binary variable for whether the household consumed ‘any fruit and vegetables from own production in the 7 days prior to the survey’. The regression analysis was conducted for a subset of women who reported buying any fruit (n=2100) and vegetables (n=2308) in the 30 days preceding the survey.

#### Fruit and vegetables intake frequency score

We ran separate multivariable linear regressions for the fruit and vegetables intake scores to examine their associations with indicators of women’s experiences in their food environment. We estimated mean differences and 95% confidence intervals (CIs) for the univariate and adjusted models.

#### Fruit and vegetables GDR variety score

We used separate Poisson regressions to estimate the association with fruit GDR score and vegetables GDR score after checking for overdispersion. We estimated risk ratios (RRs) and 95% confidence intervals (CIs) for both univariate and adjusted models, controlling for confounders.

All analyses were conducted using RStudio v 2025.5.0.496 (Posit Team, 2025). Data cleaning was done using STATA version 18 (StataCorp, 2023). Complete case analysis was used to handle missing data (<10% data missing for all variables).

### Ethical approval

Ethical approval for the study was granted by the National Institute of Medical Research in Tanzania (NIMR/HQ/R.8a/Vol.IX/4357), the International Food Policy Research Institute (IFPRI) (00007490), and Wageningen University and Research (2023-022). The data analysis for this study was approved by the University of Edinburgh (HERC_2025_047). Written consent was obtained from the household head, target woman, and retailers.

## Results

The mean (SD) age of women in our study sample was 38.2 (6.2) years. On average, 72% had attended or completed primary education, 85% were married, 59% were engaged in self-employed agriculture, and 25% were involved in non-food related activities 2 **(Table 1).** The average household size was 5.8 (1.7) members, and 49% of women reported that their household experienced severe or moderate levels of food insecurity.

**Table 1.** Demographic and household characteristics of women aged 15 to 49 years.

|  | n=2597 |
| --- | --- |
| <u>Women characteristics</u> |  |
| Age (years), M (SD) | 38.2 (6.2) |
| Highest level of education, % (n) |  |
| Never went to school | 12 (318) |
| Primary incomplete | 6 (164) |
| Primary complete | 66 (1,718) |
| Secondary incomplete | 3 (87) |
| Secondary complete | 9 (228) |
| Higher than secondary | 3 (74) |
| Primary occupation (majority of time spent), in the last 12 months, % (n) |  |
| Self-employed in agricultural activity (farming/livestock) | 59 (1,539) |
| Employed in a non-food-related activity | 25 (639) |
| Employed in food system activity | 7 (192) |
| Other (Unemployed/not seeking work/student) | 4 (109) |
| Household work, including childcare | 4 (110) |
| Marital status, % (n) |  |
| Never married | 4 (91) |
| Married or living together | 85 (2,213) |
| Widowed/divorced/separated | 11 (285) |
| <u>Household characteristics</u> |  |
| Household size, M (SD) | 5.8 (1.7) |
| Household dependency ratio, M (SD) | 1.2 (0.8) |
| Wealth quintile score, M (SD) |  |
| First/Lowest | -3.1 (0.6) |
| Second/Lower | -1.6 (0.4) |
| Third/Middle | -0.3 (0.4) |
| Fourth/Higher | 1.3 (0.5) |
| Fifth/Highest | 3.7 (1.1) |
| Household ownership of a moped, motorcycle or tricycle, % (n) | 17 (442) |
| Household ate any fruit or vegetable from their own production in the last 7 days, % (n) | 10 (271) |
| Household Food Insecurity Experience in the last 30 days, M (SD) |  |
| Any Food Insecurity (1=< FIES score <=>=8) | 63 (1,642) |
| Mild Food Insecurity (1=<FIES<=3) | 15 (384) |
| Severe or Moderate Food Insecurity (4=<FIES>=8) | 49 (1,258) |
| Abbreviations: M, mean,; SD, standard Deviation; FIES, Food Insecurity Experience Scale; |  |

Most women (81%) reported buying any fruit and any vegetables (89%) in the last 30 days **(Table 2)**. Among those who reported buying fruit and vegetables, fruit were primarily bought from either a market (80%) or kiosks (18%), and vegetables were bought from a market (40%), kiosks (27%), directly from farms (21%), or mobile vendors (11.6%). Fruit was typically bought weekly (76%), monthly (19%), or daily (5%), from the primary source. Vegetables were typically bought daily (35%) or weekly (62%). In terms of perceived availability, 65% of women reported that the desired variety of fruit was mostly available throughout the year in their primary source, and 23% reported that the desired variety of fruit was available only sometimes. For vegetables, 60% of women found the desired variety to be mostly available throughout the year, and 28% found it to be sometimes available. Two-thirds of women reported that their primary source for buying vegetables was convenient, while 65% of women reported that their primary source for buying fruit was convenient.

**Table 2:** Women’s experiences in the food environment in the previous 30 days.

|  | **Fruit**<br>**(n=2597)** | **Vegetable**<br>**(n=2597)** |
| --- | --- | --- |
| Bought any fruit or vegetable for the household, % (n) | 81 (2,114) | 89 (2,322) |
| Among those buying: |  |  |
| Primary sources of buying, % (n) |  |  |
| Market (permanent/periodic) | 80 (1,685) | 40 (925) |
| Kiosk or street stall (Genge) | 18 (384) | 27 (628) |
| Mobile vendors | 1 (19) | 12 (268) |
| Directly from the farm | 1 (14) | 21 (482) |
| Mini Market/Supermarket/Retail shop (Duka)/Other | 1 (12) | 1 (19) |
| Buying frequency from the primary source, % (n) |  |  |
| Daily | 5 (99) | 35 (823) |
| Weekly | 76 (1,612) | 62 (1,441) |
| Monthly | 19 (403) | 2 (58) |
| Availability of the desired variety throughout the year in the primary source, % (n) |  |  |
| Mostly | 65 (1,383) | 60 (1,383) |
| Sometimes | 23 (478) | 28 (644) |
| Rarely | 12.0 (253) | 13 (295) |
| Presence of market in the village for buying (Yes), % (n) | 62 (1,321) | 65 (1,515) |
| Round-trip distance between source and home in km, median (IQR) | 9.3 (2.2 - 18.6) | 3.0 (0.6 - 10.0) |
| Round-trip time between source and home in minutes, median (IQR) | 39.1 (19.4 - 78.2) | 32.0 (8.4 - 69.4) |
| Perceived convenience of buying from the primary source, % (n) |  |  |
| *Easy* | 65 (1,366) | 79 (1,842) |
| *Neutral* | 10 (210) | 5 (119) |
| *Difficult* | 25 (538) | 16 (361) |
| Primary mode of transport to the primary source, % (n) |  |  |
| Walk | 50 (1,054) | 78 (1,801) |
| Cycle | 0.4 (9) | 0.3 (7) |
| Motorbike | 21 (448) | 8 (189) |
| Public Transport | 25 (532) | 10 (232) |
| Car/Buy at home/Other | 3 (67) | 4 (90) |
| Place of origin for visiting the primary market, % (n) |  |  |
| Home | 93 (1,960) | 95 (2,204) |
| Work/Office | 7.0 (147) | 4 (96) |
| Farm | 0.0 (1) | 0.0 (1) |
| Other | 0.3 (6) | 1 (21) |

Abbreviations: M, Mean; IQR, Interquartile range (Q1 – Q3); GDR, Global Dietary Recommendation;

In the open-ended questions, women mentioned that they chose independent outlets or sellers, such as kiosks, farmers, or mobile vendors, over markets mainly because of their proximity to home (66.4%) and routine or habit (27%). In comparison, markets offered more variety (54%), better prices (40%) and were often the only source for fruit and vegetables (36%) **(supplemental table 3).**

The distance and time travelled by women to buy fruit were (Median, IQR) 9.3 km (2 - 19) and 39.1 minutes (19 - 78), while for vegetables, it were 3 km (1 - 10) and 32 minutes (8 – 69) (**Table 2)**. Walking was the primary mode of transportation for buying fruit among 50% of women and for buying vegetables among 78% of women. Motorbikes and public transport were reported as transportation modes for buying fruit among 46% of women and 18% of women for buying vegetables. Distance and time varied by sources in the food environment **(supplement figure 2).** The median (Q1 - Q3) distance and time were longest for markets (14 km (7 -24); 49 mins (33, 90)), and shortest for kiosks 1 km (0 -5); 14 mins (3, 54). Women reported that markets were inconvenient for buying fruit and vegetables due to longer travel times and greater distances **(supplement table 3).**

The most commonly consumed fruits were orange (79%), banana (69%), avocado (57%), cucumber (55%), and watermelon (53%) **(supplement table 4).** The most commonly consumed vegetables were onion (100%), tomato (99%), carrot (94%), nightshade leaves (86%), and green pepper (85%) **(supplement table 4).**. Women’s median (IQR) intake frequency score for fruit was 0.9 (0.3 - 1.8) and for vegetables was 7.5 (5.1-10.9, **Table 3).** Daily intake of any fruit was reported by 24% of women, and daily intake of any vegetables by 94% of women. Women’s daily fruit variety score was median (IQR) 0.0 (0.0-2.0), and vegetable score was 2.0 (IQR), with 15% of women reporting consuming any citrus fruit and 86% consuming any other vegetables.

**Table 3:** Women’s fruit and vegetables intake in the previous 30 days.

|  | **Fruit**<br>**(n=2597)** | **Vegetable**<br>**(n=2597)** |
| --- | --- | --- |
| Consumed any daily, n (%) | 24 (627) | 94 (2,436) |
| Consumed zero fruit or vegetables, n (%) | 6 (161) | 0% |
| Intake frequency score (out of F: 90, V: 80), Median (IQR) | 0.9 (0.3-1.8) | 7.5 (5.1-10.9) |
| Consumed any Vitamin A-rich fruit daily, n (%) | 9, (228) | - |
| Consumed any Citrus fruit daily, n (%) | 15 (392) | - |
| Consumed any Other fruit daily, n (%) | 11 (289) | - |
| Consumed any Vitamin A-rich orange vegetable daily, n (%) | - | 64 (1,667) |
| Consumed any Dark green leafy vegetable daily, n (%) | - | 36 (931) |
| Consumed any Other vegetable daily, n (%) | - | 86 (2,242) |
| Intake GDR or variety score (range 0-3), median (IQR) | 0.0 (0.0 - 2.0) | 2.0 (1.0 - 3.0) |
| Abbreviations: IQR, Interquartile range (Q1 – Q3); GDR, Global Dietary Recommendation; F, Fruit; V, Vegetables; |  |  |

In multivariable models for fruit intake frequency score, we found that women who bought fruits weekly or monthly had a lower score than women who bought daily (adjusted mean (95% CI)weekly: -1.10 (−1.50, -0.62), monthly: -1.60 (−2.00, -1.10), **(Figure 2 Panel A, and supplement table 5 Panel A).** Fruit intake frequency score was lower among women who perceived the desired variety of fruit was sometimes versus mostly available throughout the year (−0.36 (−0.51, -0.22)). Compared to the 25^th^ quintile for distance or time (<680 meters or 7 minutes, respectively), there was a positive association between median distance to the source (between 680 metres and 5.7 km) (0.27 (0.04, 0.50) and distance above the 90^th^ quintile (40 km) (0.27 (0.02, 0.51)) and fruit intake frequency score. Convenience and time were not associated with fruit intake frequency scores.

**Figure 2.**
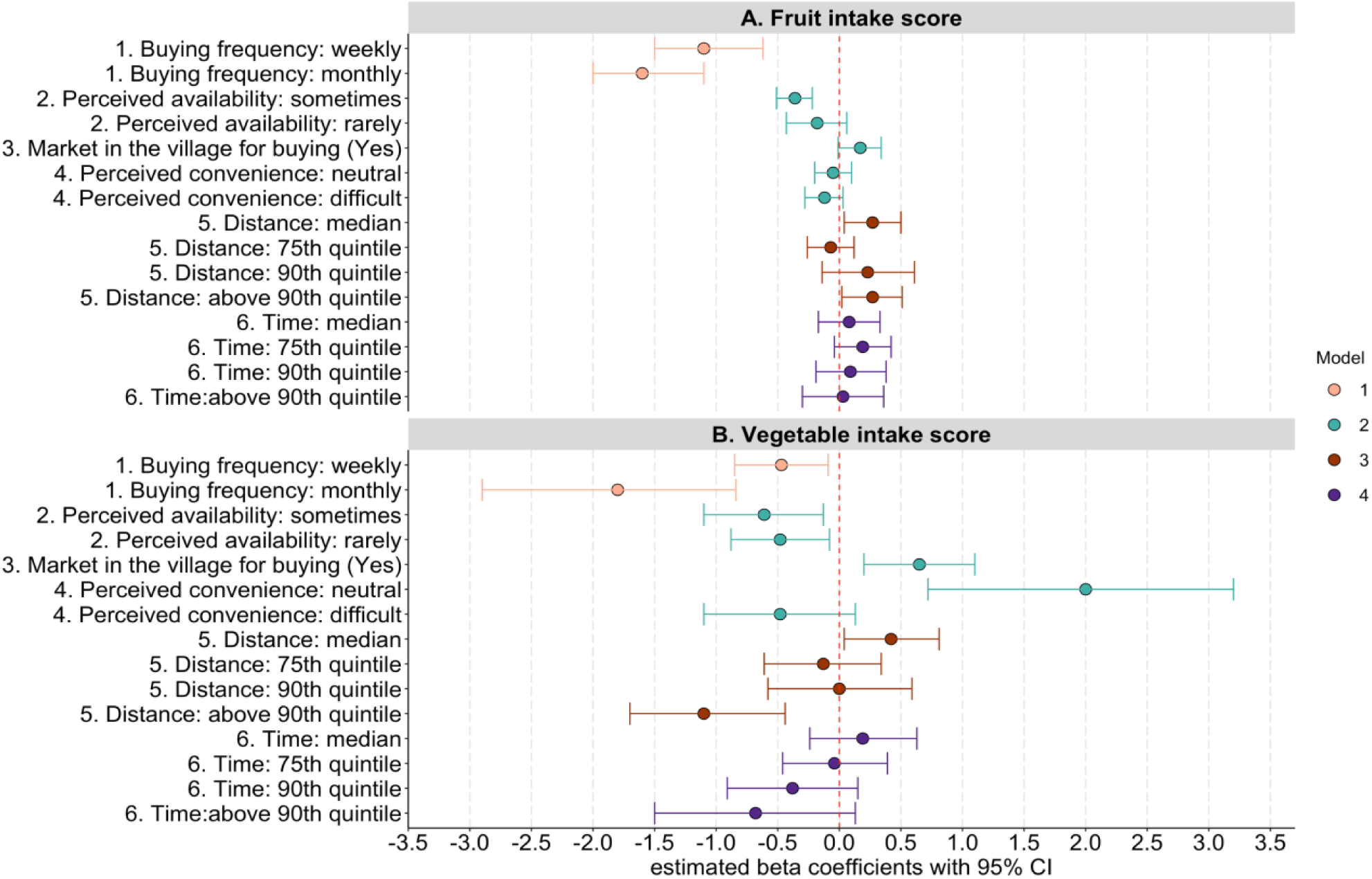
Associations of women’s food environment experiences with frequency scores of fruit and vegetables intake. Abbreviations: CI, confidence interval; Notes: Values are mean difference or relative risk and (95% CIs). The dependent variables for models in Panels A and B were frequency scores of fruit (n=2100) and vegetable (n=2308) intake, respectively, estimated using data from a non-quantitative food frequency questionnaire. The models in Panels A and B were estimated using multivariable linear regressions with standard errors accounting for clustering within the village and controlling for confounders significant at P<0.20 in univariate models. **See Supplement Table 5 for the unadjusted and full model estimates.** Model 1, exposure: buying frequency (reference: daily); covariates: household wealth, household food insecurity, women’s education, women’s occupation, own consumption, household size, women’s marital status, is included only in models in Panel B, fruit intake score. Model 2, exposures: perceived availability of desired variety throughout the year (reference: mostly), presence of market in the village, and perceived convenience (reference: easy); covariates: household wealth, household food insecurity, women’s education, women’s occupation, household size, household dependency ratio, women’s marital status is included only in models in Panel B, fruit intake score; Model 3, exposure: distance quintiles (reference: 25th quintile); covariates: household wealth, household food insecurity, own consumption, women’s education, women’s occupation, own consumption, household size and household dependency ratio, women’s marital status is included only in models in Panel B, fruit intake score; Model 4, exposure: time quintiles (reference: 25th quintile); covariates: household wealth, household food insecurity, own consumption, women’s education, women’s occupation, own consumption, household size, household dependency ratio, women’s marital status are included only in models in Panel B, fruit intake score.

Women who bought vegetables weekly and monthly had a lower intake frequency score than women who bought daily (weekly: -0.47 (−0.85, -0.09), monthly: -1.80 (−2.90, -0.84)) **(Figure 2 Panel B, and supplement table 5 Panel B**). Women who perceived the desired variety of vegetables as sometimes or rarely available throughout the year had lower vegetable intake frequency scores compared to women who perceived the desired variety of vegetables as mostly available (sometimes available: -0.61 (−1.10, -0.13); rarely available: -0.48 (−0.88, -0.08)). Women who reported the presence of a market for vegetables in the village had a higher intake frequency score (0.65 (0.20, 1.10)). Compared to the 25^th^ quintile for distance or time (less than 230 meters or 3 mins, respectively), there was a positive association between median distance (between 230 meters and 1.5 km) and vegetable intake frequency score (0.42 (0.04, 0.81)), but a negative association with distance above the 90^th^ quintile (30 km) (−1.10 (−1.70, -0.44). Convenience and time were not associated with vegetable intake frequency scores.

Women who reported buying fruit weekly or monthly, compared to daily, had a lower fruit GDR score (adjusted RR (95% CI): weekly: 0.51 (0.40, 0.64), monthly: 0.25 (0.17, 0.37), **Figure 3 Panel A, and Supplement table 6 Panel A).** Women who perceived the desired variety of fruit was sometimes available versus mostly available throughout the year also had lower fruit GDR scores (0.65 (0.51, 0.83)). Women who reported the presence of a market to buy fruit in the village had a higher variety of fruit intake (1.36 (1.14, 1.62)). We did not find a significant relationship between perceived convenience, distance or time and fruit GDR score.

**Figure 3.**
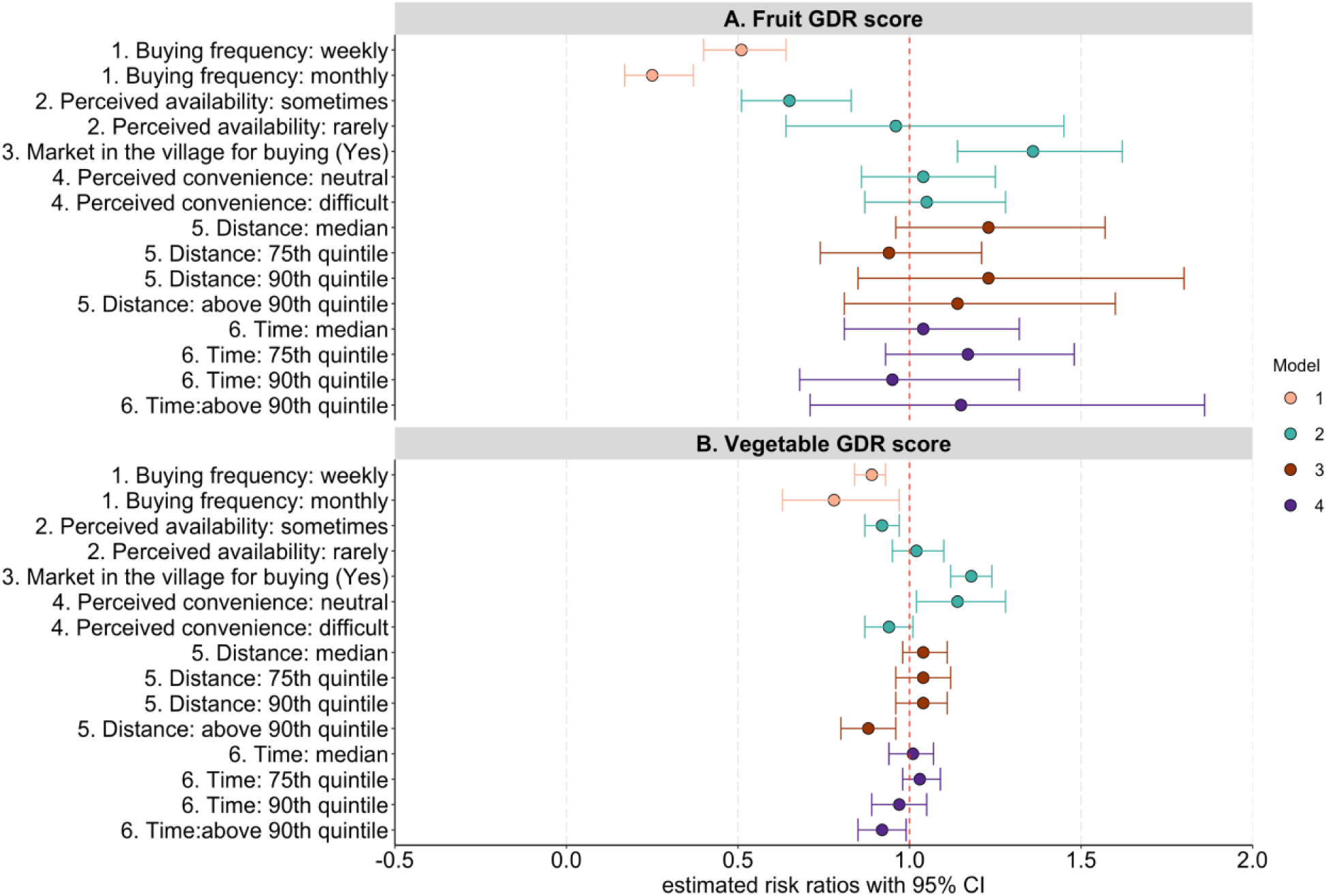
Association of food environment experiences with GDR scores of fruit and vegetables intake. Abbreviations: GDR= Global Dietary Recommendation, CI = confidence interval; Notes: Values are risk ratio estimates (95% CIs). The dependent variables for models in Panels A and B were GDR or variety scores of fruit (n=2100) and vegetable (n=2308) intake, respectively (Pastori et al., 2023), estimated using data from a non-quantitative food frequency questionnaire. The models in Panels A and B were estimated using Poisson regression with standard errors accounting for clustering within the village. The models were controlled for covariates by assessing the relationship with dependent and individual exposure variables in univariate models at P< 0.20. **See Supplement table 6 for the unadjusted and full model estimates** Model 1, exposure: buying frequency (reference: daily); covariates: household wealth, household food insecurity, own consumption, women’s education, women’s occupation, own consumption, household size; Model 2, exposures 2-4: perceived availability of desired variety throughout the year (reference: mostly), presence of market in the village, and perceived convenience (reference: easy); covariates: household wealth, household food insecurity, women’s education, women’s occupation, household size, household dependency ratio; Model 3, exposure: distance quintiles (reference: 25th quintile); covariates: household wealth, household food insecurity, own consumption, women’s education, women’s occupation, own consumption, household size and household dependency ratio; Model 4, exposure: time quintiles (reference: 25th quintile); covariates: household wealth, household food insecurity, own consumption, women’s education, women’s occupation, own consumption, household size, household dependency ratio;

Similar to fruits, women who reported buying vegetables weekly or monthly compared to daily had a lower vegetable GDR score (adjusted RR (95% CI): weekly: 0.89 (0.84, 0.93), monthly: 0.78 (0.63, 0.97), **Figure 3 Panel B, and Supplement table 6 Panel B).** Women who perceived the desired variety of vegetables was sometimes available versus mostly available throughout the year also had lower vegetable GDR scores (0.92 (0.87, 0.97)). Women who reported the presence of a market for vegetables in the village had a higher variety of vegetables in their intake (1.18 (1.12, 1.24)). A neutral perception of convenience compared to easy was associated with a higher vegetable GDR score (1.14 (1.02, 1.28)), whereas a difficult perception was not. Compared to the 25^th^ quintile for distance or time (<230 meters or 3 mins), longer distances and time above the 90^th^ quintile (>30 km or 270 minutes) were associated with a lower variety of vegetable intake (distance: 0.88 (0.80, 0.96); time: 0.92 (0.85, 0.99)).

Across all models for fruit and vegetables intake frequency and GDR scores, the relationship was positive with household wealth and negative with any household food insecurity **(Supplement tables 5 and 6).**

## Discussion

In this cross-sectional analysis of adult women from rural northern Tanzania, we found that most women reported buying both fruit and vegetables for home consumption. Vegetables were bought more frequently (daily or weekly) than fruit (weekly or monthly). Buying frequency was positively associated with both the frequency and variety of fruit and vegetable intake. Having a market in the village was associated with a higher frequency of vegetable intake and a greater variety of both fruit and vegetables intake. Women who perceived a desired variety of fruit and vegetables as less available throughout the year reported relatively lower intake frequency and variety. Vegetable sources typically used by women were more accessible than fruit sources. Longer distances were associated with a lower frequency and variety of vegetable intake.

In our study, 24% and 94% of women reported daily intake of any fruit or vegetable, respectively. This finding is consistent with findings from the Mtwara and Morogoro regions in southern Tanzania, which found that 12%-34% of women consumed fruit daily and 95% consumed vegetables daily (O’Malley et al., 2024). Similarly, another study from rural western Kenya also found that 25% of women consumed fruit daily (Keding et al., 2017). A study that examined the reasons for low fruit consumption among adults in Tanzania found that cost, education, and food beliefs, such as “fruits are regarded as snacks for children,” were important factors (Msambichaka et al., 2018). The reasons for relatively higher vegetable consumption were the women’s role in their production and preparation, as well as their lesser consumption of meals outside the home (Msambichaka et al., 2018).

Our study contributes to the emerging evidence that the majority of rural households in Africa purchase fruits and vegetables in the built food environment (Dzanku et al., 2024). A significant contribution of this paper is the use of both perceived and objective measures to quantify women’s experiences of their built food environment for fruit and vegetables, and to explore the associations with their intake of these foods. People’s perceptions of their food environment have been mostly measured for high-income countries, and studies have found the combination of perceived and objective measures to be better predictors of dietary quality and eating behaviours than objective assessment or participants’ perception alone (Gao et al., 2022; Green & Glanz, 2015; Inaç et al., 2024). While we do not have directly comparable estimates on buying frequency or perceived availability of fruit and vegetables from East Africa, one study in rural Tanzania found that having a market in the cluster (availability within a radius of 3 km from the household) had little impact on adult micronutrient intake (Ameye, 2023). In contrast, we found that women who reported having a market in the village had a higher frequency of vegetable intake and a greater variety of fruit and vegetables intake. Potential differences in results could be due to variations in study regions, and because the previous study used data that are almost 15 years old, from 2007-2008. Supporting our result, another study from rural Malawi found that having a market in the village had positive effects on individual adult diet diversity (Koppmair et al., 2017). Overall, our findings on the positive associations of buying frequency and perceived availability, with fruit and vegetables intake frequency and variety, contribute to the limited evidence on these indicators from Africa (Stadlmayr et al., 2023).

Previous studies from Africa, including rural Tanzania and rural Malawi, have found distance and time from sources (specifically, local markets) to be negatively associated with women’s dietary diversity (Keding et al., 2017; Madzorera et al., 2021; Matita et al., 2021; Nandi et al., 2021). These factors were also found to be negatively associated with household-level dietary diversity in Ethiopia and Tanzania (Usman & Haile, 2022). We have shown that distance is significant for women’s greater frequency and variety of vegetables intake and time is significant for the variety of vegetables intake. In addition, we tested the association using distance and time quintiles and did not find a dose-response relationship with intake; however, we did find a negative association between further distances and frequency and variety of vegetables intake, and a positive association between shorter distances and the frequency of both fruit and vegetables intake. Similar to our study, an earlier study from rural Tanzania found that households living within 30 minutes of a market had higher odds of purchasing nutritious foods such as dark green leafy vegetables and other Vitamin A-rich foods, and this was in turn positively associated with women’s dietary diversity (O’Malley et al., 2024).

Some of the reported differences in women’s buying experiences in the food environment between fruit and vegetables could be explained by the characteristics of the primary sources in the food environment that they accessed. Our research revealed that women primarily purchased fruit from markets and sourced vegetables from various independent outlets or sellers, including kiosks, mobile vendors, and directly from farms. The differences in source selection may be due to variations in the availability, affordability, or accessibility of fruit and vegetables between sources. A previous food environment diversity mapping exercise in 17 regional markets across Tanzania, including two markets in our study regions of Arusha and Kilimanjaro found that fruit were the second highest sold food group (in volume) in markets after grains, white roots and tubers, and plantains, while other and green leafy vegetables were fourth and six on the list (FAO, 2023). In rural Tanzania, vegetables are mainly sold in small quantities by kiosks, stationary vendors, and mobile vendors (van der Maden et al., 2021). Formal sources, such as supermarkets or formal retail outlets, represent a burgeoning trend in Tanzania, but account for less than 5% of the nation’s fresh fruit and vegetables purchases (van der Maden et al., 2021). Further research is warranted in the rural food environment to investigate the emergence of new food sources, the types of food available, and their influence on people’s food choices and diets.

Our study has several strengths. We provided evidence from rural areas on the role of consumer experiences in their food environment, contributing to the growing literature on drivers of food choices at the food group level. This study also contributes to the methodology of measuring spatial accessibility in the context of transitioning and rural food environment. First, by linking the sources in the food environment to those primarily utilised by the household, and second, by considering road path networks while accounting for the means of transport and place of origin for buying fruit and vegetables. This approach has the potential to contribute to the geographical analysis that takes into account consumers’ buying patterns and preferences in understanding the relationship between the food environment and diets (Lytle & Sokol, 2017). Given the high density and mobility of independent food sources in rural contexts, it may not be feasible to collect data on all sources and match them to household choices when examining overall diets. Therefore, the nearest sources, self-reports, and key informants, such as village or community leaders, were considered viable strategies. Indeed, these methods have been implemented by other studies on market access in Tanzania (Ameye, 2023; Madzorera et al., 2021; O’Malley et al., 2024). We further demonstrated that it is possible to enhance understanding of access beyond the analysis of geographically proximate sources by incorporating consumers’ choice of where they typically shop, especially for sources such as main markets that are central to all food purchases. Finally, while we examined both intake frequency and variety, our results on associative factors are similar for both. Future studies could consider focusing on one of these indicators.

This study also has several limitations. We used cross-sectional data; therefore, the results should be interpreted as associative rather than causal. Our outcome variables are derived from a self-reported non-quantitative fruit and vegetables FFQ, and the results are thus indicative of dietary intake and are subject to recall bias. However, non-quantitative FFQs provide relevant dietary information on the frequency of fruit and vegetables intake (Bailey, 2021), and have previously been validated among women in Tanzania (Paulo et al., 2025a; Sarfo et al., 2023; Zack et al., 2018). We further minimised misreporting and recall bias by showing photos of each fruit and vegetable during the interview. We used a simple self-reported measure of women’s convenience that considered their temporal and physical effort while buying in the built food environment. However, convenience is a multidimensional concept, and further research is needed to assess convenience within different food environments better (Bogard et al., 2024). We also recognise that fruit and vegetables intake and acquisition are seasonal. Because our data were collected during the short rainy season, our results may not reflect food experiences or consumption during the dry or long rainy seasons. A further limitation is that we did not consider the costs of fruit and vegetables, a known factor influencing fruit and vegetable consumption, especially in low-income populations (Headey et al., 2023; Stadlmayr et al., 2023). Finally, food choices and diets are driven by interactions between personal, family, cultural, food safety, and food environment factors; hence, quantitative assessments should be complemented by qualitative methods (Karanja et al., 2022; Liguori et al., 2022). Hence, we have planned future analyses, including the implementation of qualitative methods to examine the lived experiences of food environment and consumption among rural women, as well as a mixed-methods assessment of women’s food safety perceptions when buying fruit and vegetables.

## Conclusions

In conclusion, we found that a high proportion of rural women purchased fruit and vegetables in the food environment. Women’s buying frequency, perceived availability, and distance to the primary source were associated with both the frequency and variety of fruit and vegetables intake. Our findings suggest that greater availability and improved accessibility to healthier food options may lead to better dietary quality. Given the recent recommendation in Tanzania’s food-based dietary guidelines to promote daily intake of fruit and vegetables (Ministry of Health of the United Republic of Tanzania, 2023), there is a need to focus on improving the availability, affordability, and accessibility of fruit and vegetables among rural populations.

## Data Availability

All data produced in the present study can be made available upon reasonable request to the authors

## Declaration of Interest statement

The authors declare that they have no competing interests.

## Funding Sources

We would like to thank all funders who supported this research through their contributions to the CGIAR Trust Fund: www.cgiar.org/funders.

## Author contributions

The authors’ responsibilities were as follows—

Conceptualised the analyses: NS, LB, DKO, LMJ, ALB

Designed the FRESH study: LB, FA, QM, NK, SYH, JK, DKO

Contributed to data collection: NS, LB, FA, QM, EM, KJ, JK

Analysed data: NS

Wrote and contributed to the first version of the paper: NS, LMJ, ALB

Commented on subsequent versions of the paper: all the co-authors

Had primary responsibility for final content and all authors: NS, LB, LMJ, ALB

Read and approved the final manuscript: all the co-authors

## Acknowledgements

We thank all the women and households who participated in this study. We would also like to thank Gayathri Ramani, Malick Dione, Rock Zagre, and Jose Luis Torres Chaves for their support in fieldwork preparation, data management, cleaning, and analysis. We are grateful to Wiston Mwombeki for his support during fieldwork. We thank Charles D Arnold for input on the overall study design and the dietary assessment methods.

## Notes

### Competing Interest Statement

The authors have declared no competing interest.

### Author Declarations

Ethical approval for the study was granted by the National Health Research Ethics Committee, National Institute of Medical Research in Tanzania (NIMR/HQ/R.8a/Vol.IX/4357), the International Food Policy Research Institute (IFPRI) Institutional Review Board (00007490), and Wageningen University and Research's (WUR) Research Ethics Committee (2023-022). The data analysis for this study was approved by the The Royal (Dick) School of Veterinary Studies (R(D)SVS) Human Ethical Review Committee's (HERC), University of Edinburgh (HERC_2025_047). Written consent was obtained from the household head, target woman, and retailers.

