## Supplementary material for "Women’s experiences in the food environment and their association with fruit and vegetables intake: Insights from Northern Tanzania": Supplemetal file

**Article title**

**Funding Sources:** CGIAR Trust Fund: www.cgiar.org/funders.

**Supplement file** 1

Supplement figure 1: Survey locations 3

Supplement table 1: Definition of women’s experiences in the food environment variables and fruit and vegetable intake included in this study 4

Supplement table 2: Demographic variables included in the study 6

Supplement text 1: Method to calculate spatial accessibility in the food environment 8

Supplement text 2: Method to calculate women's fruit and vegetables intake score 9

Supplement text 3: Food group classification for fruit and vegetable 10

Supplement table 3: Womens’ reported reasons for selection of primary sources and for inconvenience in travel by type of reported primary sources (market versus outlets) 11

Supplement figure 2: Distance and Time measures of buying fruit and vegetables by sources in the food environments 13

Supplement table 4: Commonly consumed fruit and vegetables reported by women in the last 30 days (n=2597) 15

Supplement Table 5: Full regression tables for associations of women's food environment experiences with fruit and vegetable intake frequency scores 18

**Panel A. Dependent variable: Fruit intake frequency score (n=2100)** 18

**Panel B. Dependent variable: Vegetable Intake Score (n=2308)** 22

Supplement table 6: Association of food environment experiences with fruit and vegetable Global Dietary Recommendation (GDR) score 26

**Panel A. Dependent variable: Fruit GDR Score (n=2100)** 26

**Panel B. Dependent variable: Vegetable GDR Score (n=2308)** 29

### Supplement figure 1: Survey locations

#### Tanzania in East Africa

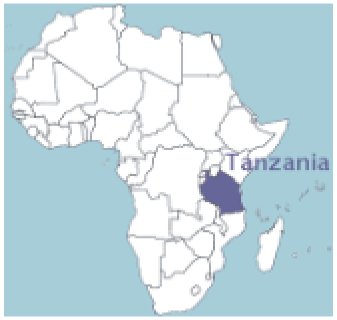

#### Study regions of Arusha and Kilimanjaro and five districts in northern Tanzania

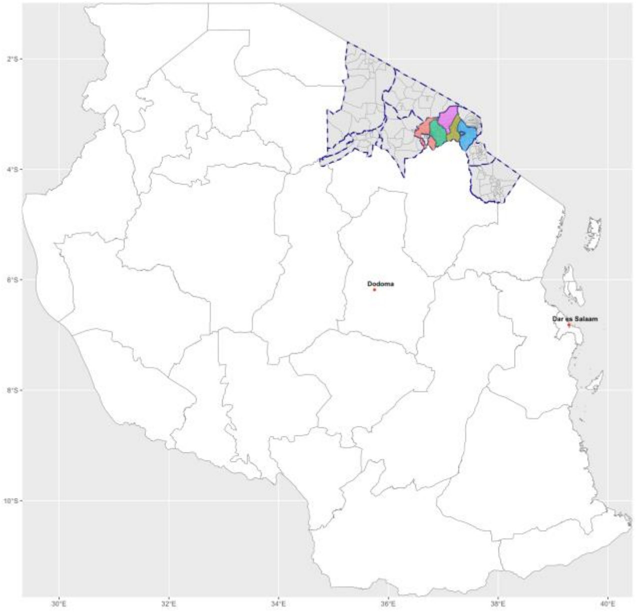

Note: Dodoma (capital city) and Dar Es Salam are key locations in Tanzania and shown here only for reference purposes. They are not study locations

#### 33 study villages in 5 Districts in northern Tanzania

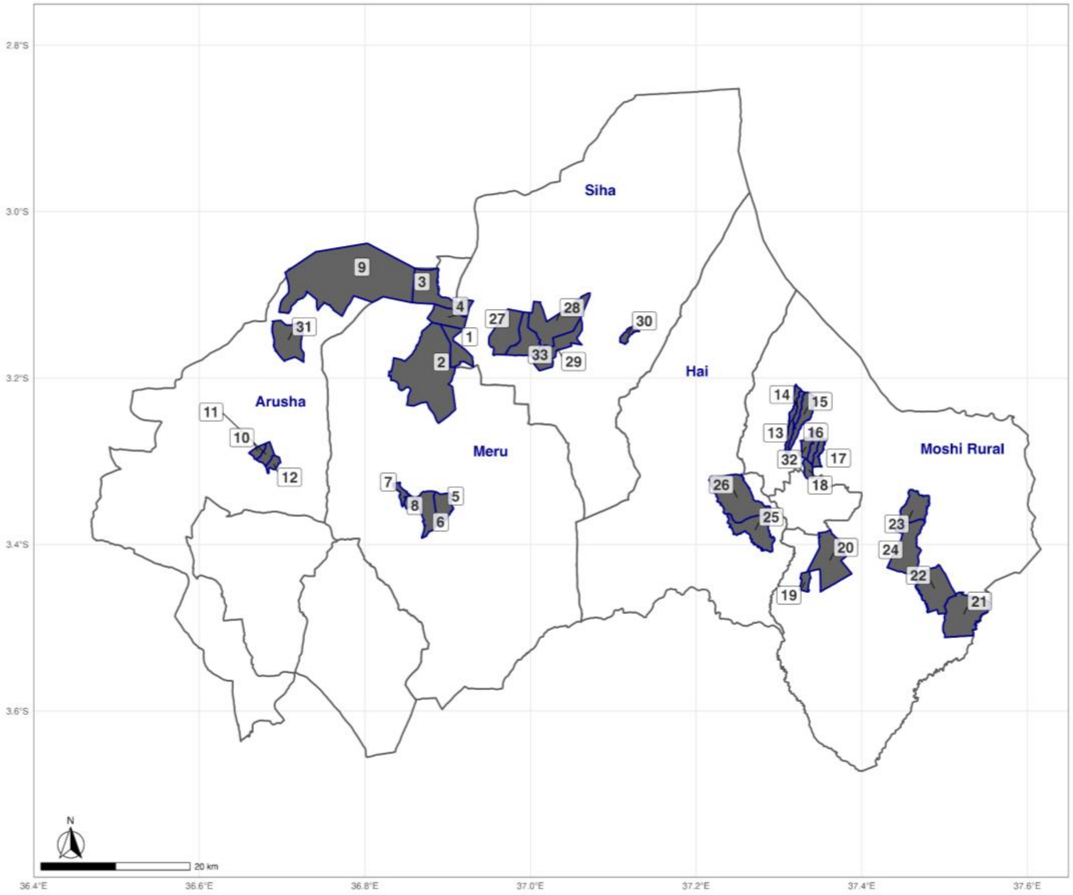

Supplement table 1: Definition of women's experiences in the food environment variables and fruit and vegetables intake included in this study

| Variable | Definition | Variable category | Measure |
|---|---|---|---|
| Sources for buying FV | Primary and secondary sources in the FE reported by women for buying FV.<br>The primary source was identified as the top-ranked source reported by women, where they typically buy most of the F&V for their household's consumption.<br>Coded: Market (periodic/permanent) (1), Kiosk or street stall or Genge (2), Mobile vendors (3), Directly from the farm (4), Minimarket/Supermarket/Retail Shop or Duka (5)<br>Market: A source operating on a daily, regular, periodic basis (e.g. weekly, fortnightly or monthly) in a designated location. Semi-permanent or permanent physical structures may be in use on market days. | Buying patterns | Self-reported behavior |
| Frequency of buying FV | Frequency of buying FV from the primary source<br>Coded: Daily (once daily, 4-6 times a week) (1), Weekly (2-3 times a week, once a week) (2), Monthly (once in 2 weeks, once a month) (3) | Buying patterns | Self-reported behavior |
| Presence of a market in the village for FV | Market in the village/town where women could buy most of the FV that the household consumes<br>Coded: No (0), Yes (1) | Buying perceptions | Self-reported behavior |
| Availability of FV throughout the year in the primary source | Primary source has a variety of FV that women want to buy throughout the year<br>Coded: Easy (1), Sometimes (1), (2) Mostly | Buying perceptions | Perceived Availability |

| | | | |
|---|---|---|---|
| Perceived convenience of buying from the primary source | Think about your commute to this [market/seller] (travel time, transport, leaving kids behind, household chores, etc.), would you consider travelling to this [market/seller] easy or difficult?<br><br>Coded: Easy (1), Neutral (2), Difficult (3) | Buying perceptions | Perceived Convenience |
| Distance | Return journey distance for FV source in FE between home and reported source conditional on primary mode of transport used (kilometres, km) | Accessibilty | Objective |
| Time | Return journey time for FV source in FE between home and reported source conditional on primary mode of transport used (in minutes); | Accessibilty | Objective |
| Primary mode of transport to the primary source | What is the primary mode of transport used to reach the [market/seller]?<br><br>Coded: Walk (1), Cycle (2), Motorbike (3), Public transport (4), Car (5) | Buying patterns | Perceived |
| Place of origin for visiting the primary market | From where do you usually commute to this [market/seller]?<br><br>Coded: Home (1), Work/Office (2), Farm (3) | Buying patterns | Perceived |
| FV intake score | Estimated intake based on weighted frequency ("2-3 times per day", "once daily", "5-6 times a day", "2-4 times a week", "once per week", "2-3 times per month", "once per month") of FV intake reported by women in the last 30 days, calculated separately for fruit and vegetables (Continuous) | Intake frequency | Estimated |
| FV GDR score | Calculated as the sum of fruit and vegetables food group consumed. The 6 FV food groups include Vitamin A-orange vegetable, Dark green leafy vegetable, Other vegetable, Vitamin A-rich fruit, Vitamin C-rich fruit, Other fruit.<br><br>Ranked data with a range of 0 to 3. | Intake variety | Estimated |

Supplement table 2: Demographic variables included in the study

| Variable | Definition |
|---|---|
| Age | Respondent women’s reported age in years (Continuous) |
| Education | Women’s Highest education attained<br>Coded: Primary incomplete (1), Primary complete (2), Secondary incomplete (3), Secondary complete (4), Higher than secondary (5) Never went to school (6) |
| Marital status | Women’s Marital Status<br>Coded: Never Married (1), Married or living together (2), Widowed or Divorced or separated (3) |
| Primary occupation | Women’s primary labor activity in the last 12 months<br><br>Coded:<br>Self-employed in agriculture activity: farming/livestock (1), Employed in non-food related activity (Enterprise/business (non-food)/Salaried government/Salaried private/non-agriculture wage labour (2),<br>Employed in food system-related activity (agriculture wage labor, food retail: market vendor/shop, kiosk, or mobile vendor food collection, transportation, or wholesale) (3),<br>Other (Unemployed/Student/Not seeking any work/ Not engaged in any activity in the last 12 months) (4),<br>Household work, including child care (5) |
| Household size | Total number of household members (count) |
| Household dependency ratio | Ratio of number of dependent household members <15 y and > 64 y to active labor force household members (15-64 y) (count) |
| Household Wealth Index quintiles | Quintiles of household wealth index created using 12 assets (radio, television, decoder ,table, armchair, burner, cabinet, wardrobe, clothes iron, fridge or freezer, shovel, moped or motorcycle or tricycle), and 7 housing characteristics (floor, roof, wall, # of rooms, electricity, clean fuel, improved toilet)household assets and other characteristics<br>Coded: Lowest (1), Lower (2), Middle (3), Higher (4), Highest (5) |

| | |
|---|---|
| Household ate any fruit and vegetables from own production in the previous 7 days | Binary indicator for any reported fruit and vegetables consumed from own cultivation in the last 7 days<br>Coded: No (0) and Yes (1) |
| Household food insecurity | Categorical indicator created using FAO's FIES (0: no insecurity to 8: severe food insecurity)<br>Coded: Any food insecurity (1, FIES >=1), Mild (2, 1=<FIES<=3), Moderate or Severe (3, FIES >=4) |

#### Supplement text 1: Method to calculate spatial accessibility in the food environment

Distance and time measures of spatial accessibility were calculated between households and sources in the food environment. We created a woman-level dataset that mapped the household geo-coordinates to their reported fruit and vegetables source in the food environment. Our approach varied based on the type of source reported for fruit and vegetables purchases because of the availability of geospatial data from the Google Maps API. For women who reported using a market as a primary source (n=2131, 48%), we were able to match the standardized market names from the household survey to the food environment data to retrieve the geo-coordinates. For the remaining women who reported using non-market outlet types as primary sources, such as kiosks or mobile vendors (n=2305, 52%), we used geospatial matching to find the nearest non-market source from home. This approach was followed due to the difficulty in matching non-market sources referred to by women to the food environment census dataset, given the non-standardized names used by women and the relatively high density of non-market sources in the food environment.

During the estimation process, we ran a couple of iterations of the code, primarily modifying the mode of transport because of the unavailability of geospatial data. In 15% of total cases (n=656), Google Maps API did not have adequate data to provide an estimate of travel time due to limited and missing data regarding public transportation. In these cases, we used estimates for driving time. This initial code modification left us with only 2% missing estimates (n=74), for whom distance was estimated over the earth's surface between two points using the Haversine formula in R (Hijmans, 2024) and time was estimated by dividing the distance over average speed for the village. The average (mean) speed at the village level was estimated for the non-missing households and then used for missing cases.

For those whose distance and/or time values were >3 SD above or below the village median value (n=66, <1%), we replaced their values with the sample village average (median). Finally, we doubled the single journey estimates to arrive at round-trip journey estimates between households and fruit and vegetables sources. While this scaling does not affect our statistical analysis, we believe it provides a closer-to-reality estimate of the time and distance involved in buying fruit and vegetables.

#### Supplement text 2: Method to calculate women's fruit and vegetables intake score

F&V intake score was calculated based on the reported frequency of intake for F&V using the following steps:

1. Intake Score = Sum of (number of F&V consumed by frequency X frequency weight)
2. Calculation of frequency weights: It is assumed that a month includes 28 days (the reference period is 4 weeks). The frequency 'one day per week' will therefore receive the value 0.14 (4/28).

**Table of frequency weights**

| Daily frequency | Weights | Scoring |
|---|---|---|
| 2-3 times per day | 2.5 | =[(((2+3)/2)*7*4)/28 ] |
| Once Daily | 1 | =[(1*7)*4/28] |
| 5-6 times a week | 0.79 | =[((5+6)/2)*4)/28] |
| 2-4 times a week | 0.43 | =[((2+4)/2)*4)/28] |
| Once per week | 0.14 | =[(1*4)/28] |
| 2-3 times per month | 0.09 | =[((2+3)/2)/28] |
| Once per month | 0.04 | =1/28 |
| Never | 0 | =0/28 |

The maximum possible score for fruit was 90 (36 items*2.5) and for vegetables it was 80 (32 items*2.5).

#### Supplement text 3: Food group classification for fruit and vegetables

Each fruit and vegetables item from the FFQ was categorized as vitamin A-rich or citrus if they were a “source” based on the Codex Alimentarius definition, i.e. 60 Retinol Activity Equivalents per 100 g for vitamin A and 9 mg per 100 g for citrus (Codex Alimentarius definition in Arimond et al., 2010). Otherwise, items were categorized based on guidelines in the FAO's Minimum Diet Diversity (MDD) for women excluding onions and spring onions that were categorized as condiment vegetables (FAO, 2021).

| Category | Items |
|---|---|
| Dark green leafy vegetables (DGLVs) | Spinach, Lettuce, Amaranth greens, Pumpkin leaves, Cowpea leaves, Sweet potato leaves, Nightshade, Cassava leaves, Spider flower, Snap beans or green beans, Ethiopian mustard, Bean leaves, Bok Choy, Jute mallow, Broccoli, Watercress, Swiss chard, Saro |
| Vitamin A-orange vegetables | Pumpkin, Carrot |
| Other vegetables (e.g., onion, garlic, spices) | Eggplant, Cabbage, Chinese cabbage, Tomato, Green pepper, Okra, Onion, African eggplant, Spring onions, Cauliflower, Mushroom, Green peas, Other vegetables |
| Vitamin A-rich fruit (ripe orange) | Mango, Papaya, Passion, Pomelo, Tree tomato, Loquat |
| Vitamin C-rich fruit (mostly citrus) | Tangerine, Lemons, Orange, Grapefruit, African Starfruit, Strychnos |
| Other fruit | Banana, Tamarind, Plums, Jackfruit, Cucumbers, Baobab, Watermelon, Guava, Peaches, Avocado, Pineapple, Bread fruits, Sour sop, Grapes, Custard apple, Cashewnut fruit, Strawberry, Dates, Pomegranate, Marula, Pears, Black plum, Kungu, Other fruit |

Supplement table 3: Womens' reported reasons for selection of primary sources and for inconvenience in travel by type of reported primary sources (market versus outlets)

| | Market | Outlets or sellers (Kiosks/retail/farms/mobile vendors) |
|---|---|---|
| **Reasons for buying from the primary source, % (n)** | N = 2,587 | N = 1,821 |
| 1. Only or convenient source | 35.9 (930.0) | 16.6 (302.0) |
| 2. Close To home or work/located on the way | 37.6 (973.0) | 66.4 (1,209.0) |
| 3. Comes Outside Our Home | 1.5 (39.0) | 14.7 (267.0) |
| 4. Best/Lowest Prices | 40.4 (1,046.0) | 31.6 (575.0) |
| 5. Personal Contact With The Vendor | 2.3 (59.0) | 7.8 (142.0) |
| 6. Offers Variety | 54.0 (1,397.0) | 31.1 (567.0) |
| 7. Offers fresh/natural product | 27.8 (719.0) | 25.2 (458.0) |
| 8. Products Have Good Appearance/Taste/Smell | 15.3 (396.0) | 15.3 (279.0) |
| 9. I Am Used To Shop/ It Is A Habit | 21.5 (557.0) | 27.1 (493.0) |
| 10. They Have Safe Products | 8.4 (218.0) | 9.4 (171.0) |
| **Reasons for inconvenience in travel to primary sources, % (n)** | N= 838 | N=53 |

| | | |
|---|---|---|
| 1. Too Far | 59.1 (495.0) | 77.4 (41.0) |
| 2. Transport Unavailable/Insufficient | 27.1 (227.0) | 7.5 (4.0) |
| 3. Takes Lot of Time | 38.1 (319.0) | 52.8 (28.0) |
| 4. No Proper Road/Infrastructure | 14.6 (122.0) | 7.5 (4.0) |
| 5. Transport cost | 2.1 (18.0) | 1.9 (1.0) |

Notes: Both these open-ended questions were asked to a subset a women who reported buying any fruit or vegetables in the last 30 days. The question on inconvenience in travel was asked to a further subset of women who reported any difficult in travel in a screening question on convenience (see table 2 in the main text for the responses to the screening question).

Supplement figure 2: Distance and Time measures of buying fruit and vegetables by sources in the food environments

**Panel A: Distance (km) by fruit and vegetables source**

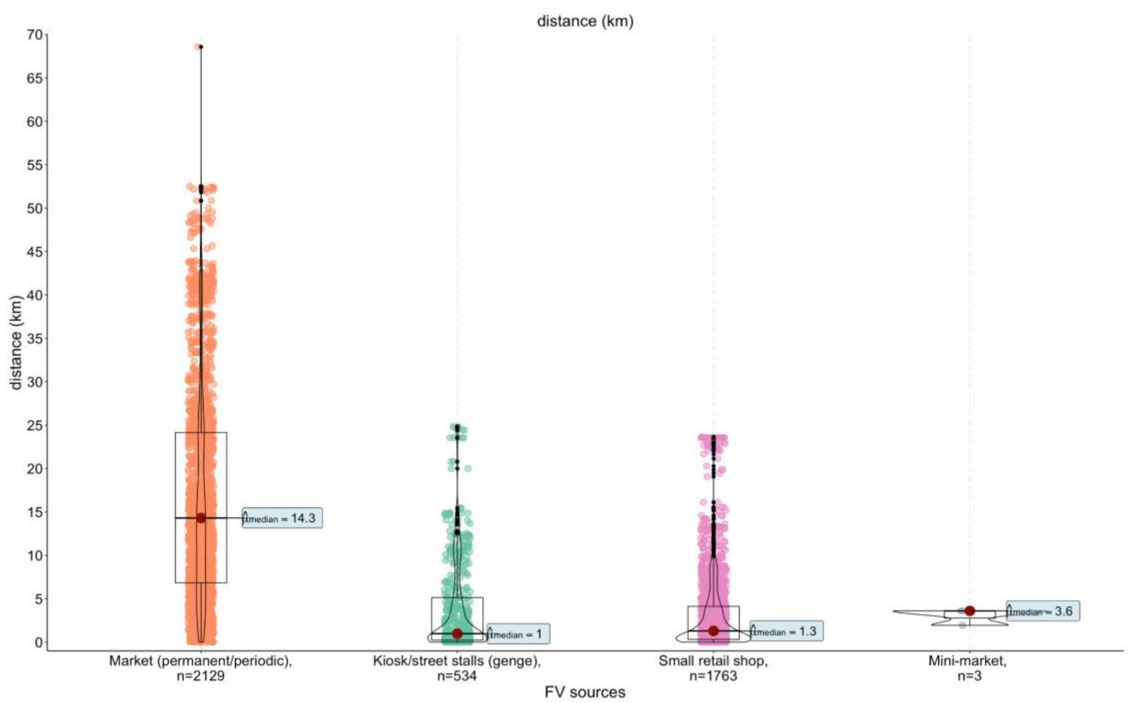

**Panel B: Time (minutes) by fruit and vegetables source i**

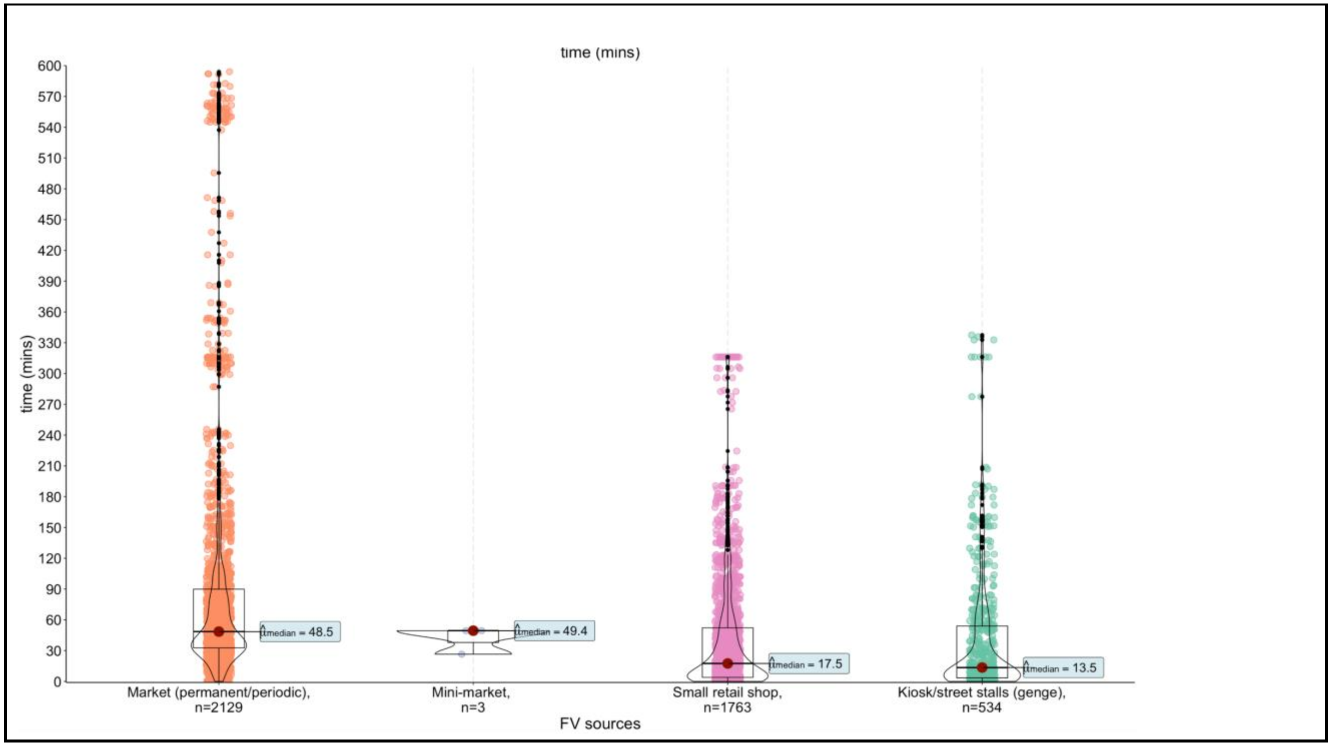

Notes: The spatial accessibility measures of distance and time were estimated for 2500 women (n=4429) using Google Maps API from home to the reported primary source or nearest source for buying fruit and vegetables and based on the mode of transport reported by women in the household survey.

Supplement table 4: Commonly consumed fruit and vegetables reported by women in the last 30 days (n=2597)

| | Fruit | % (n) |
|---|---|---|
| 1 | orange | 79% (2,063) |
| 2 | banana | 69% (1,797) |
| 3 | avocado | 57% (1,492) |
| 4 | cucumbers | 55% (1,435) |
| 5 | watermelon | 53% (1,371) |
| 6 | papaya | 49% (1,274) |
| 7 | lemons | 46% (1,184) |
| 8 | mangoes | 38% (995) |
| 9 | pineapple | 29% (763) |
| 10 | guava | 18% (480) |
| 11 | passion | 11% (283) |
| 12 | baobab | 10% (266) |
| 13 | jackfruit | 8.8% (229) |
| 14 | pomegranate | 7.9% (204) |
| 15 | tangerine | 7.8% (203) |
| 16 | grapes | 6.8% (176) |
| 17 | custard apple | 5.5% (144) |
| 18 | plums | 5.6% (146) |
| 19 | tree tomato | 6.4% (165) |
| 20 | sour sop | 5.3% (138) |
| 21 | black plum | 4.3% (113) |
| 22 | dates | 4.0% (105) |
| 23 | peaches | 4.2% (108) |
| 24 | tamarind | 4.3% (112) |
| 25 | african starfruit | 2.5% (65) |
| 26 | pears | 3.2% (84) |
| 27 | strawberry | 1.6% (42) |
| 28 | bread fruits | 0.5% (13) |

| | | |
|---|---|---|
| 29 | cashewnut fruit | 1.1% (28) |
| 30 | grapefruit | 0.8% (21) |
| 31 | kungu | 1.3% (34) |
| 32 | loquat | 0.5% (13) |
| 33 | marula | 0.3% (7) |
| 34 | pomelo | 0.4% (10) |
| 35 | stychnos | 0.3% (7) |
| | **Vegetable** | % (n) |
| 1 | onion | 100% (2,597) |
| 2 | tomato | 99% (2,574) |
| 3 | carrots | 94% (2,447) |
| 4 | nightshade leaves | 86% (2,227) |
| 5 | green pepper | 85% (2,201) |
| 6 | african eggplant | 83% (2,147) |
| 7 | amaranth green | 79% (2,049) |
| 8 | cabbage | 71% (1,835) |
| 9 | okra | 69% (1,801) |
| 10 | chinese cabbage | 63% (1,643) |
| 11 | eggplant | 61% (1,574) |
| 12 | Collard green leaves | 51% (1,314) |
| 13 | Sweet potato leaves | 48% (1,255) |
| 14 | pumpkin leaf | 47% (1,211) |
| 15 | chard | 30% (775) |
| 16 | Ethiopian mustard | 24% (630) |
| 17 | Green peas | 24% (623) |
| 18 | cowpea leaves | 19% (492) |
| 19 | spinach | 19% (490) |
| 20 | watercress | 18% (465) |
| 21 | Spider flower | 16% (412) |
| 22 | pumpkin | 13% (328) |
| 23 | Jute mallow | 12% (319) |
| 24 | green beans | 9.6% (250) |
| 25 | cassava leaves | 5.2% (136) |
| 26 | Spring onions | 4.8% (126) |
| 27 | lettuce | 4.4% (114) |

| | | |
|---|---|---|
| 28 | bean leaves | 3.5% (90) |
| 29 | broccoli | 1.3% (33) |
| 30 | cauliflower | 1.4% (37) |
| 31 | mushrooms | 0.9% (24) |

Supplement Table 5: Full regression tables for associations of women's food environment experiences with fruit and vegetables intake frequency scores

**Panel A. Dependent variable: Fruit intake frequency score (n=2100)**

| **Independent variables** | **Univariate (1)** | **(2)** | **(3)** | **(4)** | **(5)** |
|---|---|---|---|---|---|
| 1.Buying frequency | | | | | |
| Daily | Ref | — | | | |
| Weekly | -1.4 (-1.9, -0.88)*** | -1.1 (-1.5, -0.62)*** | | | |
| Monthly | -2.1 (-2.5, -1.6)*** | -1.6 (-2.0, -1.1)*** | | | |
| 2.Availability throughout the year | | | | | |
| Mostly | — | | — | | |
| Sometimes | -0.41 (-0.60, -0.21)*** | | -0.36 (-0.51, -0.22)*** | | |
| Rarely | -0.42 (-0.71, -0.13)** | | -0.18 (-0.43, 0.06) | | |
| 3.Market in the village for buying | | | | | |
| 0 | — | | — | | |
| 1 | 0.12 (-0.10, 0.34) | | 0.17 (-0.01, 0.34) | | |

| | | | | | |
|---|---|---|---|---|---|
| 4.Convenience of buying | | | | | |
| Easy | — | | — | | |
| Neutral | -0.08 (-0.25, 0.09) | | -0.05 (-0.20, 0.10) | | |
| Difficult | -0.38 (-0.58, -0.18)*** | | -0.12 (-0.28, 0.03) | | |
| 5.Distance to source quintiles | | | | | |
| 25th (680 meters) | — | | | — | |
| Median (5.7 km) | 0.11 (-0.21, 0.42) | | | 0.27 (0.04, 0.50)* | |
| 75th (13.5 km) | -0.09 (-0.45, 0.27) | | | -0.07 (-0.26, 0.12) | |
| 90th (24 km) | 0.11 (-0.41, 0.62) | | | 0.23 (-0.14, 0.61) | |
| Above 90th (40 km) | 0.17 (-0.19, 0.53) | | | 0.27 (0.02, 0.51)* | |
| 6. Time to source quintiles | | | | | |
| 25th (7 min) | — | | | | — |
| Median (30 min) | 0.07 (-0.27, 0.41) | | | | 0.08 (-0.17, 0.33) |
| 75th (55 min) | 0.12 (-0.16, 0.40) | | | | 0.19 (-0.04, 0.42) |
| 90th (101 min) | -0.12 (-0.43, 0.19) | | | | 0.09 (-0.19, 0.38) |

| | | | | | |
|---|---|---|---|---|---|
| Above 90th (290 min) | -0.42 (-0.83, -0.01)* | | | | 0.03 (-0.30, 0.36) |
| Education | | | | | |
| Primary incomplete | | — | — | — | — |
| Never went to school | | -0.05 (-0.23, 0.14) | 0.01 (-0.18, 0.21) | 0.02 (-0.18, 0.23) | 0.02 (-0.19, 0.23) |
| Primary complete | | 0.12 (-0.13, 0.38) | 0.17 (-0.08, 0.41) | 0.15 (-0.11, 0.41) | 0.17 (-0.08, 0.42) |
| Secondary incomplete | | 0.18 (-0.35, 0.70) | 0.20 (-0.32, 0.72) | 0.22 (-0.28, 0.73) | 0.24 (-0.28, 0.75) |
| Secondary complete | | 0.29 (-0.01, 0.59) | 0.33 (0.01, 0.66)* | 0.32 (0.00, 0.64) | 0.34 (0.01, 0.67)* |
| Higher | | 0.63 (0.06, 1.2)* | 0.73 (0.13, 1.3)* | 0.71 (0.10, 1.3)* | 0.75 (0.14, 1.4)* |
| Household consumed F&V from own production | | | | | |
| 0 | | — | — | — | — |
| 1 | | 0.33 (0.02, 0.64)* | 0.31 (0.00, 0.63) | 0.33 (0.01, 0.64)* | 0.32 (0.01, 0.63)* |
| Household dependency ratio | | | | -0.06 (-0.20, 0.08) | -0.06 (-0.20, 0.08) |
| Any household food insecurity | | | | | |
| No | | — | — | — | — |
| Yes | | -0.43 (-0.64, -0.21)*** | -0.49 (-0.70, -0.27)*** | -0.48 (-0.70, -0.26)*** | -0.48 (-0.69, -0.26)*** |

| | | | | | |
|---|---|---|---|---|---|
| Household size | | -0.05 (-0.09, -0.01)** | -0.05 (-0.09, -0.01)** | -0.04 (-0.08, 0.00)* | -0.04 (-0.08, 0.00)* |
| Household wealth quintiles | | | | | |
| Lowest | | — | — | — | — |
| Lower | | 0.27 (0.12, 0.42)*** | 0.25 (0.10, 0.40)** | 0.27 (0.11, 0.44)** | 0.26 (0.10, 0.42)** |
| Middle | | 0.14 (-0.04, 0.32) | 0.12 (-0.04, 0.29) | 0.15 (-0.02, 0.33) | 0.13 (-0.03, 0.30) |
| Higher | | 0.46 (0.29, 0.64)*** | 0.48 (0.32, 0.64)*** | 0.51 (0.34, 0.69)*** | 0.46 (0.29, 0.64)*** |
| Highest | | 0.71 (0.35, 1.1)*** | 0.77 (0.41, 1.1)*** | 0.81 (0.44, 1.2)*** | 0.76 (0.39, 1.1)*** |
| Marital Status | | | | | |
| Married or living together | | | — | — | — |
| Never married/Widowed/divorced/separated | | -0.30 (-0.48, -0.12)** | -0.33 (-0.52, -0.15)*** | -0.32 (-0.48, -0.16)*** | -0.31 (-0.48, -0.15)*** |
| Primary occupation | | | | | |
| Self-employed in agriculture activity (farming/livestock) | | — | — | — | — |
| Employed in non-food related activity | | 0.44 (0.23, 0.65)*** | 0.48 (0.26, 0.70)*** | 0.53 (0.33, 0.74)*** | 0.51 (0.29, 0.73)*** |
| Employed in food system activity | | 0.20 (-0.04, 0.44) | 0.18 (-0.07, 0.43) | 0.18 (-0.05, 0.42) | 0.17 (-0.08, 0.42) |
| Other (Unemployed/not seeking work/student) | | -0.16 (-0.45, 0.13) | -0.16 (-0.50, 0.18) | -0.17 (-0.50, 0.16) | -0.18 (-0.53, 0.17) |

| | | | | | |
|---|---|---|---|---|---|
| Household work including childcare | | -0.02 (-0.37, 0.34) | -0.03 (-0.38, 0.33) | -0.01 (-0.36, 0.33) | -0.02 (-0.39, 0.34) |
| No. Obs. | | | 2,100 | 2,100 | 2,100 |
| $R^2$ | | | 0.138 | 0.134 | 0.129 |
| [1] *p<0.05; **p<0.01; ***p<0.001 | | | | | |

**Panel B. Dependent variable: Vegetable Intake Score (n=2308)**

| Independent variables | Univariate (1) | (2) | (3) | (4) | (5) |
|---|---|---|---|---|---|
| 1.Buying frequency | | | | | |
| Daily | Ref | | | | |
| Weekly | -0.76 (-1.3, -0.26)** | -0.47 (-0.85, -0.09)* | | | |
| Monthly | -2.3 (-3.9, -0.84)** | -1.8 (-2.9, -0.84)*** | | | |
| 2.Availability throughout the year | | | | | |
| Mostly | Ref | | — | | |
| Sometimes | -0.72 (-1.3, -0.12)* | | -0.61 (-1.1, -0.13)* | | |
| Rarely | -1.1 (-1.6, -0.55)*** | | -0.48 (-0.88, -0.08)* | | |
| 3. Market in the village for buying | | | | | |
| 0 | Ref | | — | | |
| 1 | 0.70 (0.14, 1.3)* | | 0.65 (0.20, 1.1)** | | |
| 4. Convenience of buying | | | | | |
| Easy | Ref | | — | | |

| | | | | | |
|---|---|---|---|---|---|
| Neutral | 2.1 (0.91, 3.2)*** | | 2.0 (0.72, 3.2)** | | |
| Difficult | -1.0 (-2.2, 0.19) | | -0.48 (-1.1, 0.13) | | |
| 5.Distance to source quintiles | | | | | |
| 25th (230 meters) | Ref | | | — | |
| Median (1.5 km) | 0.18 (-0.27, 0.63) | | | 0.42 (0.04, 0.81)* | |
| 75th (5.8 km) | -0.70 (-1.2, -0.20)** | | | -0.13 (-0.61, 0.34) | |
| 90th (15 km) | -0.60 (-1.4, 0.23) | | | 0.00 (-0.58, 0.59) | |
| Above90th (30 km) | -1.7 (-2.9, -0.48)** | | | -1.1 (-1.7, -0.44)*** | |
| 6.Time to source quintiles | | | | | |
| 25th (3 mins) | Ref | | | | — |
| Median (18 min) | -0.06 (-0.64, 0.52) | | | | 0.19 (-0.24, 0.63) |
| 75th (47 min) | -0.52 (-1.1, 0.03) | | | | -0.04 (-0.46, 0.39) |
| 90th (98 min) | -0.94 (-1.5, -0.36)** | | | | -0.38 (-0.91, 0.15) |
| Above90th (270 min) | -1.6 (-2.5, -0.69)*** | | | | -0.68 (-1.5, 0.13) |
| Education | | | | | |
| Primary incomplete | Ref | — | — | — | — |
| Never went to school | | 0.14 (-0.50, 0.78) | 0.05 (-0.59, 0.68) | 0.15 (-0.51, 0.81) | 0.16 (-0.53, 0.85) |
| Primary complete | | 0.57 (-0.04, 1.2) | 0.50 (-0.10, 1.1) | 0.53 (-0.07, 1.1) | 0.59 (0.01, 1.2)* |
| Secondary incomplete | | 0.42 (-0.55, 1.4) | 0.35 (-0.67, 1.4) | 0.45 (-0.58, 1.5) | 0.46 (-0.50, 1.4) |
| Secondary complete | | 0.88 (0.32, 1.4)** | 0.87 (0.33, 1.4)** | 0.90 (0.35, 1.4)** | 0.91 (0.38, 1.4)*** |
| Higher | | 1.3 (0.30, 2.2)** | 1.2 (0.32, 2.2)** | 1.2 (0.16, 2.2)* | 1.3 (0.29, 2.2)* |
| Household consumed F&V from own production | | | | | |
| 0 | Ref | — | — | — | — |

| | | | | | |
|---|---|---|---|---|---|
| 1 | | 0.29 (-0.21, 0.79) | 0.13 (-0.35, 0.62) | 0.18 (-0.30, 0.66) | 0.18 (-0.29, 0.66) |
| Household dependency ratio | | | -0.07 (-0.29, 0.14) | -0.05 (-0.27, 0.18) | -0.04 (-0.26, 0.18) |
| Any household food insecurity | | | | | |
| No | Ref | — | — | — | — |
| Yes | | -0.94 (-1.4, -0.49)*** | -0.99 (-1.4, -0.54)*** | -0.98 (-1.4, -0.55)*** | -0.96 (-1.4, -0.53)*** |
| Household size | | 0.00 (-0.11, 0.12) | 0.00 (-0.11, 0.11) | 0.01 (-0.10, 0.12) | 0.02 (-0.10, 0.14) |
| Household wealth quintiles | | | | | |
| Lowest | Ref | — | — | — | — |
| Lower | | 1.0 (0.50, 1.5)*** | 0.93 (0.48, 1.4)*** | 0.93 (0.45, 1.4)*** | 0.99 (0.43, 1.6)*** |
| Middle | | 1.4 (0.77, 2.0)*** | 1.4 (0.82, 2.0)*** | 1.4 (0.76, 2.0)*** | 1.4 (0.69, 2.1)*** |
| Higher | | 1.6 (0.92, 2.3)*** | 1.7 (0.97, 2.4)*** | 1.6 (0.93, 2.3)*** | 1.6 (0.85, 2.3)*** |
| Highest | | 2.5 (1.9, 3.2)*** | 2.5 (1.9, 3.1)*** | 2.5 (1.8, 3.1)*** | 2.5 (1.8, 3.2)*** |
| Primary occupation | | | | | |
| Self-employed in agriculture activity (farming/livestock) | Ref | — | — | — | — |
| Employed in non-food related activity | | -0.03 (-0.43, 0.37) | -0.03 (-0.45, 0.39) | -0.03 (-0.45, 0.39) | -0.07 (-0.50, 0.37) |
| Employed in food system activity | | 0.52 (-0.21, 1.2) | 0.58 (-0.07, 1.2) | 0.46 (-0.27, 1.2) | 0.45 (-0.27, 1.2) |
| Other (Unemployed/not seeking work/student) | | -0.41 (-1.2, 0.35) | -0.30 (-1.0, 0.43) | -0.50 (-1.3, 0.26) | -0.49 (-1.2, 0.27) |
| Household work including children | | -0.06 (-1.3, 1.1) | 0.15 (-1.0, 1.3) | -0.13 (-1.3, 1.0) | -0.14 (-1.3, 1.0) |
| No. Obs. | | 2,308 | 2,308 | 2,308 | 2,308 |
| $R^2$ | | 0.108 | 0.132 | 0.111 | 0.105 |
| *p<0.05; **p<0.01; ***p<0.001 | | | | | |

Abbreviations: CI, confidence interval; Values are estimates (95% CIs). The dependent variables for models in Panel A and B were (n=2100) and vegetable (n=2308) and fruit intake frequency scores, respectively, estimated using data from a non-quantitative food frequency questionnaire. The models in Panel A and B

were estimated using multivariable linear regressions with standard errors accounting for clustering within the village and controlling for covariates by assessing relationships with dependent and individual exposure variables in univariate models at P<0.20.
Model 1, exposure: buying frequency (reference: daily); covariates: household wealth, household food insecurity, women's education, women's occupation, own consumption, household size, women's marital status is included only in models in Panel B, fruit intake score;
Model 2, exposures: perceived availability of desired variety throughout the year (ref: mostly), presence of market in the village, and perceived convenience (ref.:easy); covariates: household wealth, household food insecurity, women's education, women's occupation, household size, household dependency ratio, women's marital status is included only in models in Panel B, fruit intake score;
Model 3, exposure: distance quintiles (ref:25th quintile); covariates: household wealth, household food insecurity, own consumption, women's education, women's occupation, own consumption, household size and household dependency ratio, women's marital status is included only in models in Panel B, fruit intake score;
Model 4, exposure: time quintiles (ref:25th quintile); covariates: household wealth, household food insecurity, own consumption, women's education, women's occupation, own consumption, household size, household dependency ratio, women's marital status is included only in models in Panel B, fruit intake score;
We also checked for variance inflation factor (vif) values to test for multicollinearity between covariates and most of them had values between 1 and 1.5 suggesting low multicollinearity.

Supplement table 6: Association of food environment experiences with fruit and vegetables Global Dietary Recommendation (GDR) score

**Panel A. Dependent variable: Fruit GDR Score (n=2100)**

| Independent variables | Univariate (1) | (2) | (3) | (4) | (5) |
|---|---|---|---|---|---|
| 1.Buying frequency | | | | | |
| Daily | — | — | | | |
| Weekly | 0.44 (0.34, 0.59)*** | 0.51 (0.40, 0.64)*** | | | |
| Monthly | 0.18 (0.13, 0.27)*** | 0.25 (0.17, 0.37)*** | | | |
| 2.Availability throughout the year | | | | | |
| Mostly | — | | — | | |
| Sometimes | 0.59 (0.45, 0.77)*** | | 0.65 (0.51, 0.83)*** | | |
| Rarely | 0.75 (0.49, 1.14) | | 0.96 (0.64, 1.45) | | |
| 3.Market in the village for buying | | | | | |
| 0 | — | | — | | |
| 1 | 1.33 (1.07, 1.65)** | | 1.36 (1.14, 1.62)*** | | |

| | | | | | |
|---|---|---|---|---|---|
| 4.Convenience of buying | | | | | |
| Easy | — | | — | | |
| Neutral | 1.01 (0.83, 1.23) | | 1.04 (0.86, 1.25) | | |
| Difficult | 0.86 (0.66, 1.11) | | 1.05 (0.87, 1.28) | | |
| 5.Distance to source quintiles | | | | | |
| 25th | — | | | — | |
| Median | 1.11 (0.83, 1.48) | | | 1.23 (0.96, 1.57) | |
| 75th | 0.95 (0.69, 1.31) | | | 0.94 (0.74, 1.21) | |
| 90th | 1.15 (0.73, 1.79) | | | 1.23 (0.85, 1.80) | |
| Above90th | 1.11 (0.78, 1.57) | | | 1.14 (0.81, 1.60) | |
| 6.Time to source quintiles | | | | | |
| 25th | — | | | | — |
| Median | 1.04 (0.77, 1.40) | | | | 1.04 (0.81, 1.32) |
| 75th | 1.14 (0.88, 1.47) | | | | 1.17 (0.93, 1.48) |
| 90th | 0.85 (0.62, 1.16) | | | | 0.95 (0.68, 1.32) |

| | | | | | |
|---|---|---|---|---|---|
| Above90th | 0.84 (0.51, 1.38) | | | | 1.15 (0.71, 1.86) |
| Education | | | | | |
| Primary incomplete | | — | — | — | — |
| Never went to school | | 1.13 (0.70, 1.83) | 1.22 (0.78, 1.91) | 1.24 (0.77, 1.98) | 1.22 (0.76, 1.96) |
| Primary complete | | 1.35 (0.78, 2.35) | 1.44 (0.84, 2.47) | 1.43 (0.82, 2.49) | 1.44 (0.84, 2.47) |
| Secondary incomplete | | 1.67 (0.80, 3.46) | 1.72 (0.84, 3.50) | 1.76 (0.88, 3.50) | 1.78 (0.88, 3.59) |
| Secondary complete | | 1.43 (0.80, 2.55) | 1.57 (0.88, 2.82) | 1.56 (0.86, 2.83) | 1.58 (0.87, 2.86) |
| Higher | | 1.18 (0.62, 2.26) | 1.35 (0.70, 2.59) | 1.29 (0.67, 2.49) | 1.34 (0.71, 2.55) |
| Household consumed F&V from own production | | | | | |
| 0 | | — | — | — | — |
| 1 | | 1.18 (0.94, 1.48) | 1.15 (0.91, 1.44) | 1.19 (0.94, 1.50) | 1.19 (0.95, 1.48) |
| Household dependency ratio | | | 0.90 (0.77, 1.05) | 0.90 (0.77, 1.06) | 0.90 (0.77, 1.06) |
| Any household food insecurity | | | | | |
| No | | — | — | — | — |
| Yes | | 0.68 (0.53, 0.86)** | 0.62 (0.48, 0.79)*** | 0.64 (0.50, 0.82)*** | 0.64 (0.50, 0.83)*** |

| | | | | | |
|---|---|---|---|---|---|
| Household size | | 0.96 (0.90, 1.03) | 0.96 (0.90, 1.03) | 0.97 (0.91, 1.04) | 0.97 (0.90, 1.04) |
| Household wealth quintiles | | | | | |
| Lowest | | — | — | — | — |
| Lower | | 1.35 (0.93, 1.95) | 1.31 (0.91, 1.90) | 1.35 (0.91, 2.01) | 1.36 (0.92, 2.00) |
| Middle | | 1.27 (0.84, 1.91) | 1.26 (0.84, 1.90) | 1.29 (0.84, 1.97) | 1.26 (0.84, 1.91) |
| Higher | | 1.65 (1.08, 2.53)* | 1.69 (1.11, 2.57)* | 1.71 (1.10, 2.65)* | 1.66 (1.06, 2.58)* |

**Panel B. Dependent variable: Vegetable GDR Score (n=2308)**

| Independent variables | Univariate (1) | (2) | (3) | (4) | (5) |
|---|---|---|---|---|---|
| 1. Buying frequency | | | | | |
| Daily | — | — | | | |
| Weekly | 0.87 (0.82, 0.92)*** | 0.89 (0.84, 0.93)*** | | | |
| Monthly | 0.76 (0.58, 0.98)* | 0.78 (0.63, 0.97)* | | | |
| 2. Availability throughout the year | | | | | |
| Mostly | — | | — | | |
| Sometimes | 0.88 (0.82, 0.94)*** | | 0.92 (0.87, 0.97)** | | |

| | | | | |
|---|---|---|---|---|
| Rarely | 0.94 (0.86, 1.03) | | 1.02 (0.95, 1.10) | |
| 3. Market in the village for buying | | | | |
| 0 | — | | — | |
| 1 | 1.19 (1.12, 1.27)*** | | 1.18 (1.12, 1.24)*** | |
| 4. Convenience of buying | | | | |
| Easy | — | | — | |
| Neutral | 1.16 (1.04, 1.30)** | | 1.14 (1.02, 1.28)* | |
| Difficult | 0.91 (0.80, 1.03) | | 0.94 (0.87, 1.01) | |
| 5. Distance to source quintiles | | | | |
| 25th | — | | | — |
| Median | 1.02 (0.95, 1.10) | | | 1.04 (0.97, 1.10) |
| 75th | 0.98 (0.90, 1.07) | | | 1.04 (0.97, 1.12) |
| 90th | 0.99 (0.90, 1.08) | | | 1.04 (0.96, 1.11) |
| Above 90th | 0.83 (0.74, 0.94)** | | | 0.88 (0.80, 0.96)** |
| 6. Time to source quintiles | | | | |

| | | | | | |
|---|---|---|---|---|---|
| 25th | — | | | | — |
| Median | 0.99 (0.92, 1.07) | | | | 1.01 (0.94, 1.07) |
| 75th | 0.99 (0.93, 1.07) | | | | 1.03 (0.98, 1.09) |
| 90th | 0.92 (0.84, 1.00) | | | | 0.96 (0.89, 1.05) |
| Above 90th | 0.86 (0.78, 0.94)*** | | | | 0.92 (0.85, 1.00)* |
| Education | | | | | |
| Primary incomplete | | — | — | — | — |
| Never went to school | | 1.06 (0.94, 1.19) | 1.05 (0.94, 1.17) | 1.06 (0.95, 1.19) | 1.06 (0.94, 1.20) |
| Primary complete | | 1.10 (0.98, 1.23) | 1.09 (0.97, 1.23) | 1.10 (0.97, 1.24) | 1.10 (0.98, 1.24) |
| Secondary incomplete | | 1.07 (0.94, 1.23) | 1.05 (0.91, 1.21) | 1.07 (0.93, 1.24) | 1.07 (0.93, 1.24) |
| Secondary complete | | 1.14 (1.02, 1.28)* | 1.14 (1.02, 1.28)* | 1.14 (1.02, 1.29)* | 1.14 (1.02, 1.29)* |
| Higher | | 1.16 (1.00, 1.34) | 1.14 (0.98, 1.32) | 1.14 (0.99, 1.32) | 1.15 (0.99, 1.33) |
| Household consumed F&V from own production | | | | | |
| 0 | | — | — | — | — |

| | | | | | |
|---|---|---|---|---|---|
| 1 | | 1.08 (1.00, 1.16) | 1.05 (0.97, 1.13) | 1.06 (0.98, 1.14) | 1.06 (0.98, 1.14) |
| Household dependency ratio | | | 1.00 (0.97, 1.02) | 1.00 (0.97, 1.03) | 1.00 (0.97, 1.03) |
| Any household food insecurity | | | | | |
| No | | — | — | — | — |
| Yes | | 0.92 (0.88, 0.97)** | 0.91 (0.87, 0.96)*** | 0.92 (0.88, 0.97)** | 0.92 (0.88, 0.97)** |
| Household size | | 1.01 (1.00, 1.03) | 1.01 (1.00, 1.03) | 1.01 (1.00, 1.03) | 1.02 (1.00, 1.03) |
| Household wealth quintiles | | | | | |
| Lowest | | — | — | — | — |
| Lower | | 1.14 (1.07, 1.22)*** | 1.13 (1.07, 1.19)*** | 1.14 (1.07, 1.21)*** | 1.14 (1.07, 1.23)*** |
| Middle | | 1.15 (1.05, 1.25)** | 1.16 (1.07, 1.26)*** | 1.16 (1.07, 1.27)*** | 1.16 (1.06, 1.27)** |
| Higher | | 1.21 (1.10, 1.33)*** | 1.23 (1.13, 1.35)*** | 1.23 (1.12, 1.35)*** | 1.22 (1.10, 1.35)*** |
| Highest | | 1.19 (1.08, 1.30)*** | 1.21 (1.11, 1.31)*** | 1.21 (1.11, 1.33)*** | 1.21 (1.09, 1.33)*** |
| Primary occupation | | | | | |
| Self-employed in agriculture activity (farming/livestock) | | — | — | — | — |

| | | | | | |
|---|---|---|---|---|---|
| Employed in non-food related activity | | 1.03 (0.96, 1.10) | 1.03 (0.96, 1.11) | 1.04 (0.98, 1.11) | 1.03 (0.97, 1.10) |
| Employed in food system activity | | 1.02 (0.94, 1.10) | 1.03 (0.96, 1.11) | 1.02 (0.95, 1.10) | 1.02 (0.94, 1.10) |
| Other (Unemployed/not seeking work/student) | | 0.99 (0.90, 1.10) | 1.02 (0.93, 1.12) | 1.00 (0.91, 1.10) | 1.0 (0.90, 1.10) |
| Household work including children | | 1.11 (1.00, 1.22)* | 1.13 (1.01, 1.27)* | 1.11 (1.01, 1.23)* | 1.10 (1.00, 1.22) |
| No. Obs. | | 2,308 | 2,308 | 2,308 | 2,308 |
| Notes: CI, confidence interval; Values are estimates (95% CIs) | | | | | |
| 1 *p<0.05; **p<0.01; ***p<0.001 | | | | | |
| 2 IRR = Incidence Rate Ratio | | | | | |

**Notes:** Abbreviations: GDR= Global Dietary Recommendation**,** CI, confidence interval; Values are risk ratio estimates (95% CIs). The dependent variable for models in Panel A and B were fruit (n=2100) and vegetable (n=2308) GDR score respectively (Pastori et al., 2023), estimated using data from a non quantitative food frequency questionnaire. The models in Panel A and B were estimated using Poisson regression with standard errors accounting for clustering within the village. The models were controlled for covariates by assessing relationship with dependent and individual exposure variables in univariate models at P<0.20.covariates.

Model 1, exposure: buying frequency (reference: daily); covariates: household wealth, household food insecurity, own consumption, women's education, women's occupation, own consumption, household size;

Model 2, exposures 2-4: perceived availability of desired variety throughout the year (ref: mostly), presence of market in the village, and perceived convenience (reference: easy); covariates: household wealth, household food insecurity, women's education, women's occupation, household size, household dependency ratio;

Model 3, exposure: distance quintiles (reference:25th quintile); covariates: household wealth, household food insecurity, own consumption, women's education, women's occupation, own consumption, household size and household dependency ratio;

Model 4, exposure: time quintiles (ref:25th quintile); covariates: household wealth, household food insecurity, own consumption, women's education, women's occupation, own consumption, household size, household dependency ratio;

We also checked for variance inflation factor (vif) values to test for multicollinearity between covariates and most of them had values between 1 and 1.5 suggesting low multicollinearity.
